# Evaluating primary and booster vaccination prioritization strategies for COVID-19 by age and high-contact employment status using data from contact surveys

**DOI:** 10.1101/2022.12.02.22283040

**Authors:** Ethan Roubenoff, Dennis Feehan, Ayesha S. Mahmud

## Abstract

The debate around vaccine prioritization for COVID-19 has revolved around balancing the benefits from: (1) the direct protection conferred by the vaccine amongst those at highest risk of severe disease outcomes, and (2) the indirect protection through vaccinating those that are at highest risk of being infected and of transmitting the virus. While adults aged 65+ are at highest risk for severe disease and death from COVID-19, essential service and other in-person workers with greater rates of contact may be at higher risk of acquiring and transmitting SARS-CoV-2. Unfortunately, there have been relatively little data available to understand heterogeneity in contact rates and risk across these demographic groups. Here, we retrospectively analyze and evaluate vaccination prioritization strategies by age and worker status. We use a mathematical model of SARS-CoV-2 transmission and uniquely detailed contact data collected as part of the Berkeley Interpersonal Contact Survey to evaluate five vaccination prioritization strategies: (1) prioritizing only adults over age 65, (2) prioritizing only high-contact workers, (3) splitting prioritization between adults 65+ and high-contact workers, (4) tiered prioritization of adults over age 65 followed by high-contact workers, and (5) tiered prioritization of high-contact workers followed by adults. We find that for the primary two-dose vaccination schedule, assuming 70% uptake, a tiered roll-out that first prioritizes adults 65+ averts the most deaths (31% fewer deaths compared to a no-vaccination scenario) while a tiered roll-out that prioritizes high contact workers averts the most number of clinical infections (14% fewer clinical infections compared to a no-vaccination scenario). We also consider prioritization strategies for booster doses during a subsequent outbreak of a hypothetical new SARS-CoV-2 variant. We find that a tiered roll-out that prioritizes adults 65+ for booster doses consistently averts the most deaths, and it may also avert the most number of clinical cases depending on the epidemiology of the SARS-CoV-2 variant and the vaccine efficacy.

## 1 Introduction

COVID-19 vaccines have been shown to be highly effective at preventing severe illness and death (Baden et al. 2021; Polack et al. 2020). Following the introduction of vaccination in the U.S. in December 2020, infection rates decreased dramatically through the first quarter of 2021 as increasing shares of the population were protected via vaccine-derived immunity (Gupta et al. 2021). Due to a limited vaccine supply initially, there has been a complicated debate around the trade offs of vaccine prioritization strategies for adults over age 65, healthcare workers, other essential frontline workers, and the general public (Schaffer DeRoo, Pudalov, and Fu 2020; Persad, Peek, and Emanuel 2020; Persad, Emanuel, et al. 2021; Giubilini, Savulescu, and Wilkinson 2021).

Control interventions for infectious diseases can have different public health objectives: while the highest priority is often to limit total deaths, secondary priorities can include limiting total infections or reducing the number of infected persons at a given time to below a critical care capacity threshold (Bubar et al. 2021; Buckner, Chowell, and Springborn 2021). Prioritization of vaccination towards a specific group has the direct benefit of reducing infections and deaths in that group. However, for vaccines that prevent transmission, there is also an indirect benefit of limiting secondary infections. As such, the total effect of vaccination—the sum of the direct and indirect effects on incidence and deaths—extends beyond the benefits conferred to recipients of vaccination themselves.

The risk of severe disease, hospitalization, and death from COVID-19 increases sharply with age (Levin et al. 2020; O’Driscoll et al. 2021), indicating that vaccinating adults 65+ may be most effective at reducing total hospitalizations and deaths due to COVID-19. On the other hand, in-person workers with higher rates of person-to-person contacts are at an increased risk of being infected with and transmitting SARS-CoV-2. Thus, vaccine prioritization strategies need to balance: (1) the direct protection conferred by the vaccine amongst those at highest risk of severe disease outcomes, and (2) the benefits of indirect protection and potentially achieving herd immunity more quickly through vaccinating those that are at highest risk of being infected and of transmitting the virus. In theory, the indirect benefits of preventing COVID-19 by prioritizing high-contact workers could outweigh the direct effects of vaccinating adults 65+ (Buckner, Chowell, and Springborn 2021).

Vaccinating high-contact workers also has important implications for social and economic equity. Historically-disadvantaged groups, especially Blacks and Hispanics, are over-represented in essential and front-line occupations and have younger age-distributions compared to Whites Nelson et al. 2022. Prioritizing high-contact workers, therefore, delivers proportionally more doses to racial and ethnic groups that have also been hardest hit by the pandemic (Andrasfay and Goldman 2021; Wrigley-Field, Garcia, et al. 2020; Wrigley-Field, Kiang, et al. 2021) compared to a purely age-based prioritization.

In the US, distribution of the primary vaccine doses prioritized a combination of adults 65+ and essential workers in progressive phases, beginning with those living in long-term care settings and healthcare workers. Eligibility was first opened to adults 65+ and to certain occupational groups before opening to the general public (Dooling 2021). The debate on prioritization is, however, still relevant in many other parts of the world, as well as in the US for future booster doses. It is also important to retrospectively evaluate prioritization strategies to inform response to future pandemics. Mathematical models that account for both the direct and indirect effects of vaccination can help guide policy decisions on prioritization. However, there is little data on contact rates by worker status and age available for the U.S., making these models hard to parameterize. The relatively few contact surveys conducted during 2020-2021 indicated that total contacts had substantially reduced compared to pre-pandemic measures (C. Y. Liu et al. 2021, D. M. Feehan and Mahmud 2021). However, previously available data has reported contact rates disaggregated by age, but not by both age and occupational status. A nationally representative study reported that most contacts during this period did happen at work, and that nonwhite and workers in essential occupations did have among the highest total contact rates Nelson et al. 2022. Here, we retrospectively analyze and evaluate vaccination prioritization strategies by age and worker status using detailed contact data from surveys to parameterize a mathematical transmission model for SARS-CoV-2.

We compare total effects of prioritizing adults 65+ for vaccination versus prioritizing workers who potentially have a higher risk of contracting COVID-19 due to their in-person work status (Baker, Peckham, and Seixas 2020; Hawkins 2020; Selden and Berdahl 2020). The model accounts for contact patterns between age and occupation groups using contact survey data collected as part of the Berkeley Interpersonal Contact Survey (BICS; D. M. Feehan and Mahmud 2021), which has been collecting detailed information about a respondent’s daily behavior and their disease-relevant interpersonal contact since March 2020.

We find that prioritizing adults 65+ for primary vaccination averts 25% more deaths than prioritizing high contact workers and 11% more than a split strategy. However, the most clinical infections are averted by strategies that prioritize high contact workers first.

We extend the model to consider the prioritization of booster doses during a hypothetical future outbreak of a new SARS-CoV-2 variant that is able to partially evade vaccine-derived immunity from primary vaccination. When considering booster doses, the reduction in deaths is greatest when prioritizing adults 65+, but all three strategies have similar effects on clinical infections. These results highlight the impact of various vaccination prioritization strategies and can help guide policies during future outbreaks.

## 2 Methods

### 2.1 Data

The BICS survey, collected in several waves beginning in March 2020, is an online survey aimed at capturing the frequency and nature of respondents’ physical and conversational contacts over a 24-hour period. Respondents (egos) in the BICS survey are asked about members of their household and their total number of non-household contacts, as well as detailed information on up to 3 of their previous day’s contacts (alters). Here we use data from wave 4 of the BICS survey, collected between November 30th and December 8th, 2020, where respondents were asked additional questions on their work status and work contacts. Respondents who reported being employed were asked to indicate the number of close contacts they had while performing duties for their job.

Respondents are divided into 3 categories: low contact (LC) adults (age 18-64) not reporting having any close work contacts, high contact (HC) adults (age 18-64) reporting having work contacts, and adults 65+. 34% of respondents aged 18-64 reported having interpersonal contacts at work and are labeled as high contact (HC). Children (under age 18) were not included in the survey, although adult respondents could report their contact with children. The procedure for estimating contacts for the 0-18 age group is described below. Alters are divided into children aged 0-18, adults 18-64, and adults 65+ and weighted by the number of total close contacts reported by each ego (see D. M. Feehan and Mahmud 2021 for more detail on the weighting procedure). Working-age alters aged 18-64 are categorized as being work contacts if the reported relationship was a coworker or client or if the reported contact happened at work or a store. This consisted of 22% of reported contacts for adults aged 18-64. For some working-age alters we were unable to determine work status from the survey data. This was due to missing data for non-household alters on the purpose of the reported contact and for all reported household contacts (since their work status was not collected). For the alters whose in-person work status is indeterminable from the survey data provided, alters are randomly re-labeled as having in-person work such that the proportion of high contact alters matched the surveyed ego proportion. We also perform a sensitivity test to understand the impact of this random reallocation (see supplementary section S1.3).

The survey responses are used to construct an age and work-status structured contact matrix between children, low-contact adults (18-64), high-contact adults (18-64), and adults 65+ (Figure 1) using standard methods described in more detail by D. M. Feehan and Mahmud 2021 and Jarvis et al. 2020. Briefly, the raw contact matrix ***M*** has entries *m*_*ij*_ corresponding to the average number of daily contacts between respondents (ego) in group *i* with their reported contacts in group *j*, adjusted for survey weights. Total contacts in a population must be reciprocal, but may not be in the survey data due to sampling and differences in survey reporting. We impose reciprocity using previously described methods (D. M. Feehan and Mahmud 2021). To adjust for reciprocity, population data for each group is taken from the American Community Survey (ACS) 2019 5-year estimates (US Census Bureau 2019). Adults aged 18-64 in the ACS are categorized as working in-person (34%) and not working in-person (66%) using the survey proportions since equivalent data on in-person work status is unavailable from the ACS. As children are not included in the BICS survey, contacts between children are derived from the POLYMOD survey as described in the supplementary materials section S1.2. The reciprocal contact matrix, ***C***, is the reciprocity-enforced average daily contact matrix.

**Figure 1:**
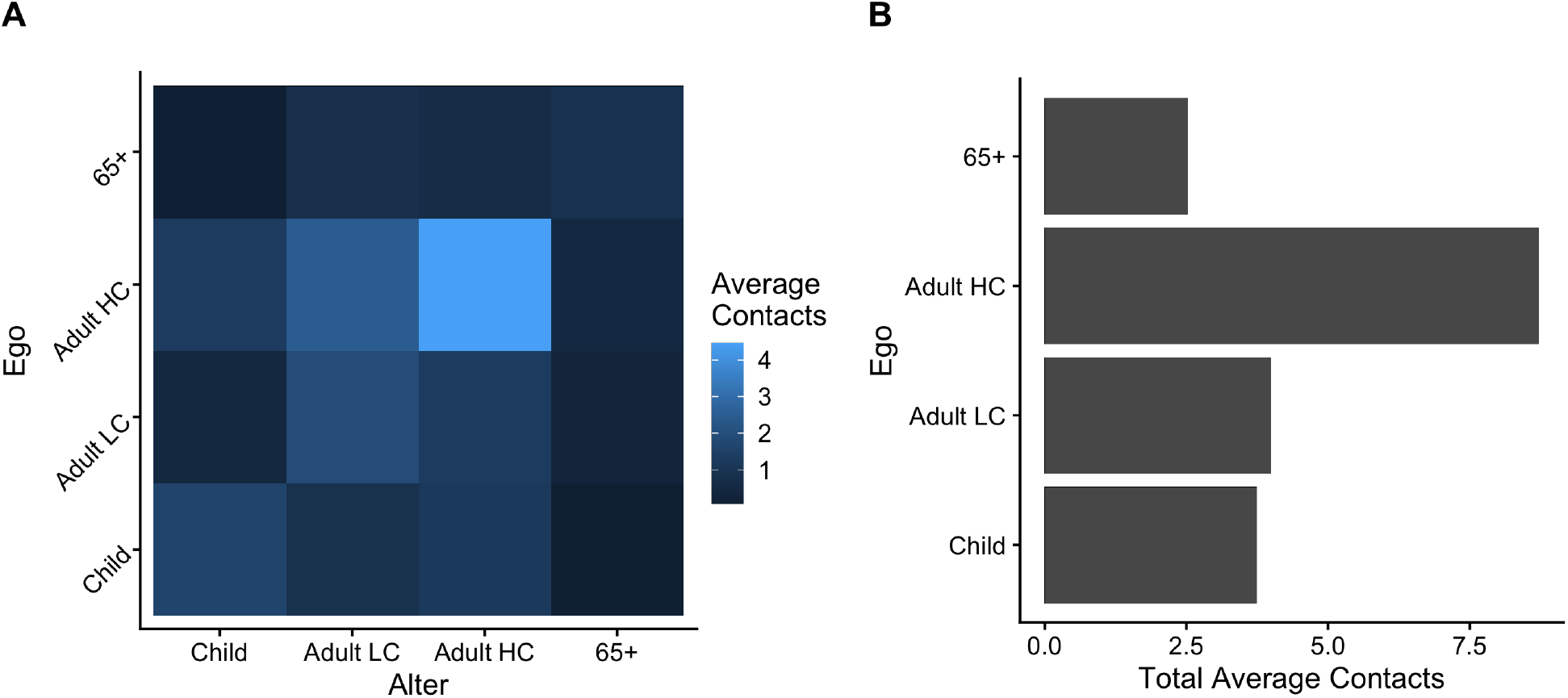
(A) Age and work-status structured contact matrix showing daily average number of reported contacts, after adjusting for reciprocity in total contacts and survey weights. (B) Sum of average number of reported daily contacts for each group. For both figures, “Adult LC” and “Adult HC” correspond to adults without and with in-person work contacts (Low Contact and High Contact, respectively). Within-group contacts for children (0-18) are derived from the POLYMOD survey (Mossong et al. 2008).

### 2.2 SARS-CoV-2 transmission model

To model SARS-CoV-2 transmission dynamics and COVID-19 incidence and mortality, we use a deterministic, continuous time compartmental model (outlined in the supplementary section S1.5, similar to Bubar et al. 2021 and Buckner, Chowell, and Springborn 2021. The model allows for heterogenous mixing between age and in-person employment status groups as specified by the contact matrix derived from the BICS survey data. Susceptibles (compartments *S* and *S*_*x*_, described below) who are exposed to SARS-CoV-2 enter an exposed (latent) phase (*E*). Depending on their age group, exposed individuals proceed to have clinical (symptomatic) infection (*I*_*c*_) with probability [0.35, 0.4, 0.75] for children, adults 18-64, and adults 65+ respectively; remaining cases experience subclinical (asymptomatic) infection (*I*_*sc*_; adapted from Davies et al. 2020). We allow for subclinical transmission, at a reduced probability (50%; Davies et al. 2020) relative to clinical infections. Probability of death after clinical infection (*μ*) varies according to age; subclinical mortality is assumed to be zero. Fatal cases proceed to compartment *D* and non-fatal cases recover to compartment *R*.

We model a two-dose primary vaccination schedule, distributed 25 days apart, to match the two-dose Pfizer and Moderna vaccines that account for the majority of the vaccination doses delivered in the United States (CDC 2020). Susceptibles awaiting the vaccine in compartment *S* proceed to compartment *V*_*a*_ after the first dose and then to *V*_*b*_ after the second dose; individuals who have contracted SARS-CoV-2 are ineligible for the vaccine. We incorporate ‘leaky’ vaccine efficacy and vaccine hesitancy using methods similar to Bubar et al. 2021 and assume an 80% reduction in infections after the first dosage and 90% after the second, consistent with efficacy estimates during the initial roll-out of the vaccines. (Tenforde 2021; Thompson 2021). Breakthrough infections occurring among those who have received either the first or second dose proceed to the exposed compartment and then on to the infected compartments (with the same probabilities as unvaccinated exposed individuals). Vaccine hesitancy is incorporated by imposing a 70% uptake of the primary vaccination doses, derived from the National Immunization Survey’s May 2021 primary uptake for seniors (National Center for Immunization and Respiratory Diseases (NCIRD) 2022b; National Center for Immunization and Respiratory Diseases (NCIRD) 2022a); the remaining 30% of susceptibles who refuse or are otherwise ineligible for the vaccine are placed in the compartment *S*_*x*_, and are otherwise identical to individuals in the *S* compartment. At its peak, nearly 5 million Americans were being vaccinated per day (CDC 2020); however, as initially vaccine rollout was slower, we assume an average 2 million vaccinations per day distributed equally between first and second shots. First doses are distributed until the *S* compartment is depleted, either through vaccination or infection; after distribution of vaccines to the priority group, vaccines are distributed to any remaining adults 65+, adults 18-64, and children proportional to the remaining susceptible population size in each group. For simplicity, those who have recovered from infection are not eligible for vaccination in our simulations.

The next generation matrix (NGM; Bansal, Grenfell, and Meyers 2007; Diekmann, Heesterbeek, and Roberts 2010; Bubar et al. 2021) for the model is:

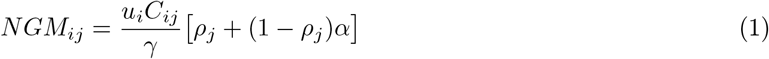

Where *γ* is the recovery rate, *ρ* is the probability that an exposed individual becomes clinically infectious, *u*_*i*_ is the age-dependent susceptibility to infection after contact with an infectious individual, and *C*_*ij*_ is the entry in contact matrix ***C*** corresponding to the average number of daily contacts a respondent in group *i* has with an individual in group *j*. The dominant eigenvalue of the NGM is the basic reproduction number, *R*_0_. We scaled the values of *u*_*i*_ to calibrate to a specific value of *R*_0_ by optimizing a scaling factor for *u*_*i*_ such that the largest eigenvalue of the NGM matches an assumed *R*_0_ value (Davies et al. 2020; Bubar et al. 2021).

The starting population for the simulations is the United States population on January 1st, 2021, with approximately 73.4 million children, 200.5 million adults 18-64, and 50.8 million adults 65+ (US Census Bureau 2019). Working-age adults were split into working in person and not working in person (including unemployed) using the survey proportions of 34% and 66%, respectively, as derived from BICS data. On January 1st, there were 20, 166, 028 confirmed cases, 352, 390 deaths, and 11, 426, 602 known active cases. The starting conditions for deaths, subclinical cases, recovered cases, and exposed individuals are detailed in section S1.

### 2.3 Model parameters

Estimates for *u*_*i*_ and *ρ*_*i*_ are taken from literature (Davies et al. 2020; Bubar et al. 2021). To account for uncertainty in other model parameters, we performed 1000 simulations with transmission and mortality parameters drawn from their assumed distributions (described below) using latin hypercube sampling. *R*_0_ was assumed to be distributed normally with mean 2.5 and standard deviation 0.54 (following D. M. Feehan and Mahmud 2021). Child, adult 18-64, and adult 65+ mortality were drawn from uniform distributions [0.003%, 0.005%], [0.20%, 0.26%], [6.9%, 10.4%], respectively (bounds for uniform distribution are the 95% confidence intervals from Levin et al. 2020 for age groups 0-34, 45-54, and 75-84). Average latent period is assumed to be distributed uniformly between 2 and 4 days and average duration of infectiousness is assumed to be distributed uniformly between 4 and 6 days, such that median draws follow values assumed by the literature (see supplemental table S1.8 for sources on all parameters). Additional parameters are outlined in table S1.8. For all simulations, we estimated the percent reduction in total clinical infections and total deaths from January 1, 2021 until December 31st, 2021 compared to a no-vaccination scenario for five vaccine prioritization strategies: (1) prioritizing only adults over age 65, (2) prioritizing only adults 18-64 with in-person work contacts, (3) splitting priority vaccines evenly between adults 65+ and adults working in person, (4) a ‘tiered’ strategy that prioritizes adults 65+ before high contact workers, and (5) a ‘tiered’ strategy that prioritizes high contact workers before adults 65+ (further details are provided in the supplementary material S1.7). The ‘tiered’ strategies are intended to replicate the CDC’s decision to progressively distribute the vaccine in a series of decreasing priorities. In our simulations, tiered roll-outs differ from the single priority strategies by allowing a second-priority group to have access to the vaccine before general distribution (when doses are distributed proportionally to eligible group size). For example, for the single priority 65+ strategy, after all of those eligible in the oldest age group have been vaccinated, remaining vaccines are distributed to other groups proportional to remaining eligible group size; however, during the tiered 65+ strategy, after eligible adults 65+ have been vaccinated doses are distributed to eligible HC adults before being distributed to other groups. For clarity, we present the simulation results using the median draw for each parameter for discussion of effect sizes (*R*_0_ = 2.5, *μ* = [0.00004, 0.0023, 0.08], latent period = 3 and infections period = 5), but show the full range of simulation results across the 1000 parameter combinations.

### 2.4 Booster dose model for a subsequent outbreak of a SARS-CoV-2 variant

We also extend the model to consider a subsequent outbreak caused by a new, more transmissible SARS-CoV-2 variant, such as the Omicron variant, where vaccines may be less effective (Collie et al. 2022; Hogan et al. 2021; UK Health Security Agency 2021; Thompson 2022). In this situation, distribution of a third “booster” dose is necessary to increase protection against clinical infection, hospitalization, and death. To maintain consistency across results, the starting population size is the same as in the previous simulation (the US population on January 1st 2021). In the booster simulation, we take the January 1st 2022 estimate of 34%, 78%, and 95% of children, adults 18-64, and adults 65+ as having received a primary course of vaccination (2 doses, represented by compartment *V*_*b*_; National Center for Immunization and Respiratory Diseases (NCIRD) 2022a; National Center for Immunization and Respiratory Diseases (NCIRD) 2022b). Remaining susceptibles are assumed to refuse the vaccine and are placed in compartment *S*_*x*_. We assume that 70% of individuals in *V*_*b*_ who have received the two-dose primary course of vaccination will receive booster doses; hesitancy or ineligibility for booster doses is incorporated by moving 30% of these individuals in *V*_*b*_ to *V*_*bx*_, indicating that they decline the booster dose. One million booster doses are distributed daily among individuals in *V*_*b*_ until that compartment reaches zero individuals either through infection or vaccination. In this booster model, no individuals will be present in compartment *S* (awaiting first dose) or *V*_*a*_ (awaiting awaiting second dose). We consider the same five strategies as before for priority distribution of booster doses. Since the vaccine’s effectiveness in reducing transmission is unknown for primary and boosted individuals for new variants, we conduct 1000 simulations with randomly drawn values for vaccine efficacy from an assumed distribution; we also draw 1000 values of *R*_0_ from an assumed distribution. The vaccine efficacy is randomly drawn from distributions derived from the CDC’s estimates of vaccine efficacy against the Omicron variant (Thompson 2022). In these simulations, primary (2-dose) vaccine efficacy is drawn from a uniform distribution between 32% and 43%; vaccine efficacy of the booster dose is drawn from a uniform distribution between 79% and 84%. (Thompson 2022). Breakthrough infections among individuals who have received 2 or 3 doses of the vaccine proceed through the Exposed and Infectious compartments. A meta-analysis of the Omicron variant’s *R*_0_ estimates shows considerable variation from 5.5 to 24, with a median of 10 (IQR: 7.25, 11.88; Y. Liu and Rocklöv 2022). To account for a wide range of transmissibility of potential new variants, we draw *R*_0_ uniformly between 2 and 12. All other parameters are held at their median values as above. Similar to the primary simulation, we present results from a simulation with the median draw of all parameters.

## 3 Results

### 3.1 Prioritization strategies for primary vaccine doses

In the simulation with median parameter values, tiered 65+ roll out reduces deaths by 31.32% (723866 deaths averted) compared to a no-vaccination scenario. This strategy saves 25.13% (532,567) more lives than prioritizing high contact workers, and 11.49% (206,087) more lives than splitting prioritization between workers and adults 65+. We note that this strategy is only marginally more effective at reducing deaths (0.34%) than prioritizing only adults 65+ before general distribution, indicating that any distribution strategy that gives initial priority to older adults will limit the most deaths. For a tiered 65+ roll out, there is also a modest reduction in clinical infections— 13% fewer (10.9 million infections averted) compared to no vaccination. However, we find that the optimal strategy for limiting clinical infections is through a tiered roll-out that first prioritizes high contact workers. This strategy reduces infections by 13.9% compared to no vaccination (11.5 million clinical infections averted), although we note that the reduction in clinical infections is similar among all prioritization schemes. Overall, we find that strategies that prioritize high contact workers, even when split with adults 65+, do limit clinical infections but fail to confer the lifesaving benefit of strategies prioritizing adults over age 65.

Figure 2a shows the percent reduction in clinical deaths and infections relative to no vaccination, across a range of values for the mortality and transmission parameters. Our results are consistent across a wide range of parameter combinations. In 100% of the simulations, the tiered 65+ roll-out was the optimal strategy for limiting deaths due to COVID-19. All prioritization strategies performed remarkably similarly for reducing the most number of clinical infections. In 62.5% of simulations the tiered HC roll-out was the most effective at limiting clinical infections; in the remaining 37.5% of simulations, the most effective strategy appears to be the tiered 65+ roll-out. Through sensitivity analysis, we show below that these differences are driven by variations in *R*_0_.

**Figure 2:**
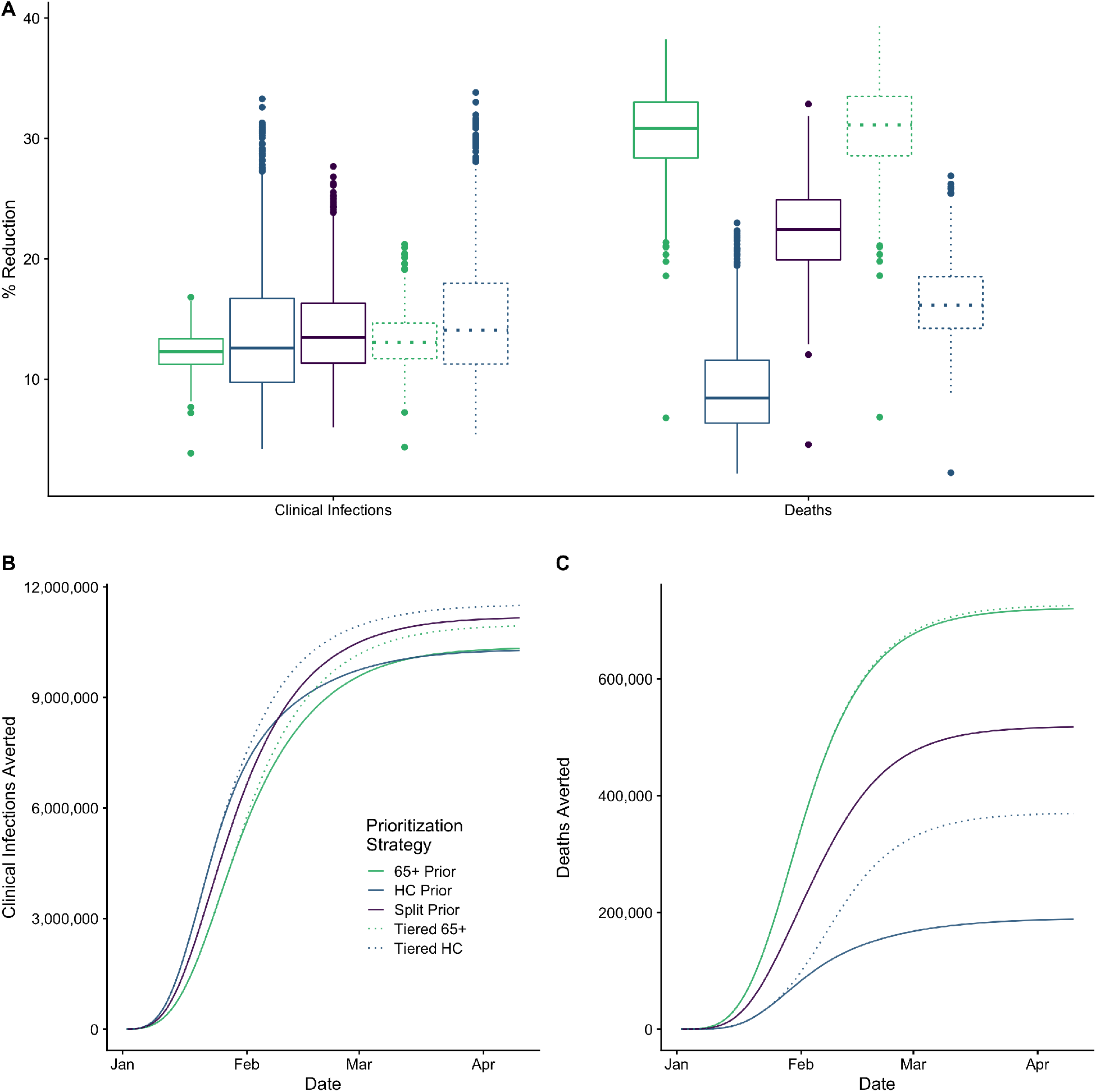
(A) For primary vaccination, percent reduction in clinical infections and deaths when compared to no vaccination for randomly drawn transmission parameters. The median percent reduction in deaths was highest in a tiered strategy that prioritizing seniors and lowest when only prioritizing contact risk workers; clinical infections are reduced the most by a tiered system that prioritizes workers first, although all strategies produce similar results. (B) and (C): For baseline parameters, trajectories of daily cumulative clinical infections (B) and deaths (C) averted relative to a no vaccination scenario, calculated as the cumulative difference between each strategy and null through each date. When prioritizing seniors the reduction in deaths begins nearly immediately, whereas the indirect benefit from prioritizing HC workers begins later and is lower in magnitude. The opposite is observed for clinical infections.

Trajectories of cumulative clinical infections and deaths averted for the five prioritization strategies with median parameter values (figure 2b, 2c) indicate a marked departure from no vaccination between mid January and February. Vaccines reach their peak lifesaving power very quickly within the first month and the relative benefit increases through February. This demonstrates that the timing of vaccines is critical: prioritizing seniors limits the most deaths because they are able to develop vaccine-derived immunity before the peak of the outbreak. Further, tiered roll out strategies show an extended benefit over their single-prioritization counterparts through February and March after distribution of vaccines to the first priority group. Trajectories for each demographic group are shown in the model appendix. Interestingly, when deaths are broken down by age and worker status, the tiered 65+ strategy averts the most deaths only in the adults 65+ group. For all other groups (children 0-18 and adults 18-64), the tiered HC strategy averts the most deaths. However, since deaths are relatively much higher in the adults 65+ group, the tiered 65+ strategy averts the most deaths in the population overall (see Supp Fig S4).

We conduct additional analyses to test the sensitivity of our results to the choice of simulation parameters. For each simulation, we separately vary *R*_0_ between 1 and 5, *μ*_65+_ between 1% and 10%, and the proportion of priority vaccinations split between high contact workers and adults 65+ between 0 and 1. All other parameters are kept the same as the baseline. We see that for all values of *R*_0_, the deaths are lowest by prioritizing adults 65+ for vaccination (figure 3). However, the strategy for averting the most clinical infections is sensitive to *R*_0_. A cross-over in optimal strategy to limit clinical infections occurs around *R*_0_ = 2.6. When *R*_0_ is less than 2.6, prioritizing high contact workers results in the fewest clinical infections. With higher values of *R*_0_, prioritizing adults 65+ leads to the fewest clinical infections. With larger values of *R*_0_, the peak of the outbreak happens earlier, resulting in more cases before most vaccines are distributed. Since high-contact individuals in a population will be infected earlier in an outbreak (Mossong et al. 2008) and, therefore, removed from the eligible pool of vaccine recipients, the strategies prioritizing high-contact workers are no longer optimal for reducing clinical infections for high values of *R*_0_. Instead, for high values of *R*_0_ prioritizing adults 65+ is optimal both for reducing deaths as well as clinical infections. However, these results are dependent on the timing of vaccine introduction and the practicalities of reaching a large enough population for vaccination before the peak of the outbreak.

**Figure 3:**
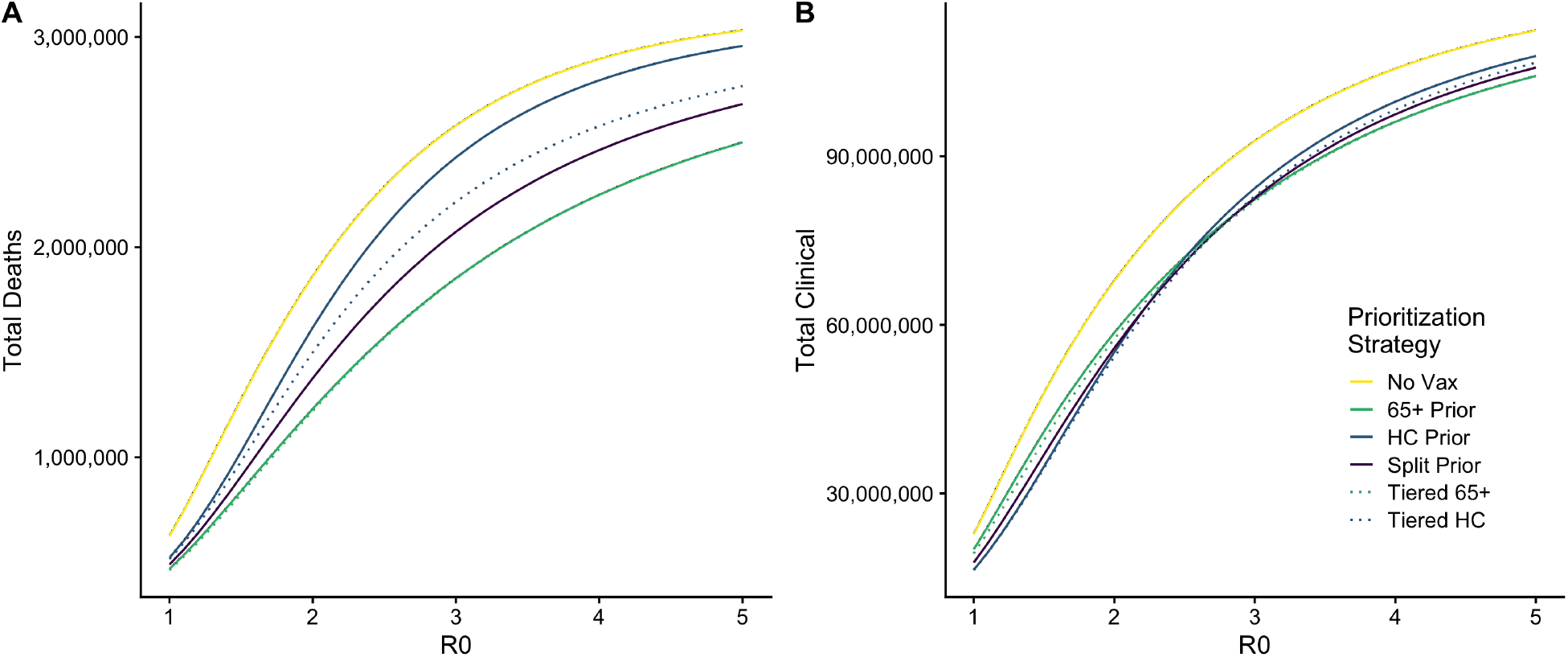
Results of the sensitivity analysis of *R*_0_ (varied linearly between 1 and 5 in 0.1 increments) and the resultant count of total deaths (A) and clinical infections (B). Across all values, a strategy that prioritizes adults 65+ either directly or in a tiered roll out limits the most deaths. At low values of R0 (<2.5) the reduction in clinical infections is greatest in a ‘Tiered HC’ roll out; however, the most effective strategy with high R0 is through a Tiered 65+ strategy.

As expected, the reduction in the number of deaths is highly sensitive to *μ* (supplemental figure S7); however, the most effective strategy consistently remains prioritizing adults 65+ for vaccination due to the strong age gradient in mortality. Figure 4 shows the effect on total deaths and clinical infections as the proportion of vaccinations given to adults 65+ is varied under the split vaccination scenario. Deaths are always minimized when giving 100% of priority vaccines to adults 65+, and total infections are minimized when priority is given to high contact workers. However, we do find a minimum number of clinical infections occurring when 55% of vaccinations are given to seniors. This non-linearity in clinical infections is a result of the fact that the burden of clinical infections by subgroup is jointly determined by the age-dependent susceptibility to infection (higher amongst HC workers) and probability of symptomatic illness (highest amongst adults 65+).

**Figure 4:**
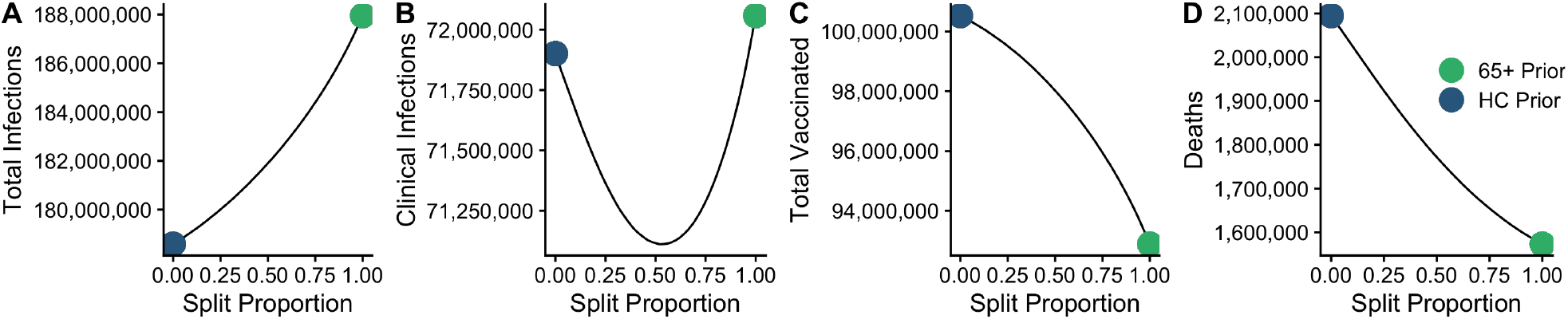
The proportion of vaccines split between seniors and HC workers is varied from 0% to 100% and shown with counts of (A) total infections, (B) clinical infections, (C) total vaccinated, and (D) total deaths. Extremes (corresponding to the HC Prior and 65+ Prior strategies) are shown. When priority vaccines are given more to adults 65+, deaths are minimized but total infections are maximized, indicating that while this strategy limits deaths it fails to limit transmission effectively. Additionally, more susceptibles are eligible for vaccination under this strategy. However, the minimum number of clinical infections is minimized when 59% of vaccines are distributed to 65+. This effect is mediated by increased susceptibility to infection and increased probability of symptomatic infection among seniors, and the increased priority group size among HC workers.

### 3.2 Prioritization of Booster Doses during a subsequent outbreak

In the simulation conducted with the median parameter draws, prioritization has a smaller effect on relative outcomes for booster doses than the primary doses. A tiered 65+ roll out for boosters not only continues to save the most lives, but in a departure from the primary simulation, also reduces clinical infections the most—although like the primary simulation, the reduction in infections is nearly equal between the three strategies. With median parameter values (*R*_0_: 7; primary vaccine efficacy: 37.5%; booster vaccine efficacy: 81.5%), a tiered 65+ roll-out reduces deaths by 14.23% (393,562) compared to no booster doses. This strategy reduces deaths by 11.5% more than prioritizing only HC workers and 7% more than a tiered HC roll out. Tiered 65+ roll-out reduces infections by 5.8% (6.5 million infections averted); however, all three strategies reduce infections by nearly the same amount (within 1%). We note that in this subsequent outbreak, the population of eligible HC workers awaiting vaccination is considerably smaller than the primary outbreak; while the eligible 65+ population between the two scenarios is similar (33 million adults 65+ are awaiting primary vaccination in the primary outbreak compared to 31 million for booster doses), the population of eligible high contact workers decreases from about 45 million to about 35 million. This is driven by lower uptake rates of primary vaccination among those below age 65 compared to adults 65+. These results are replicated across the 1000 sets of simulated parameters, shown in figure 5. Across all 1000 simulations the median percentage reduction in deaths compared to no boosters is greatest during a tiered 65+ roll out. Clinical infections are reduced the most during a tiered 65+ roll out in 49.1% of simulated parameters; in the other 50.9% of simulations, tiered HC roll out was most effective. Similarly to the primary scenario, this variation in the number of clinical infections averted is driven by *R*_0_.

**Figure 5:**
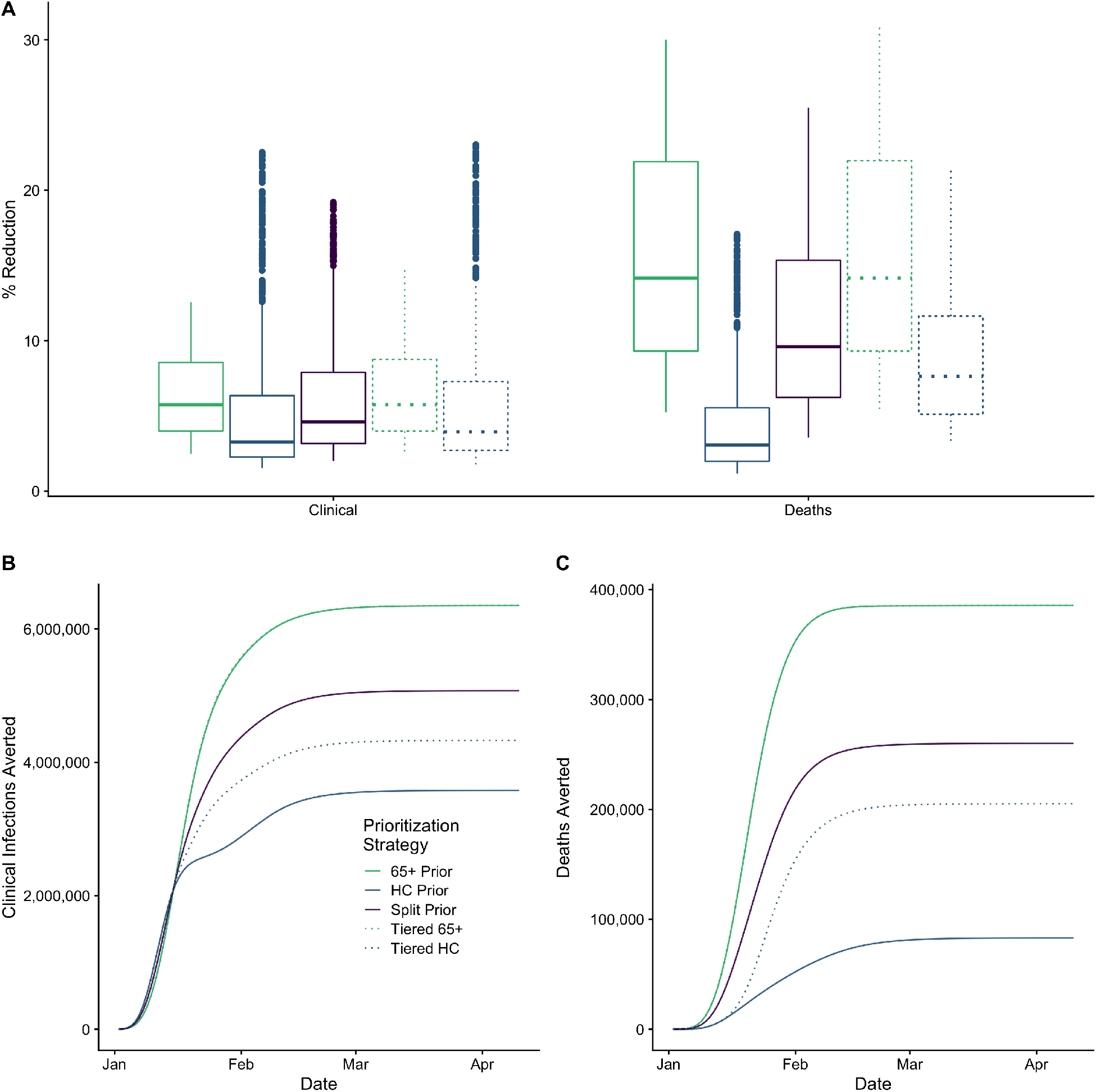
For distribution of booster doses: (A) percent reduction in clinical infections and deaths when compared to no vaccination for randomly drawn transmission parameters. (B) and (C): For baseline parameters, counts of clinical infections (B) and deaths (C) averted relative to a no vaccination scenario.

When *R*_0_ is low, HC or Tiered HC prioritization can limit the most clinical infections (figure 6). For both deaths and clinical infections the difference between all distribution strategies is smaller when *R*_0_ is higher (figure 6).

**Figure 6:**
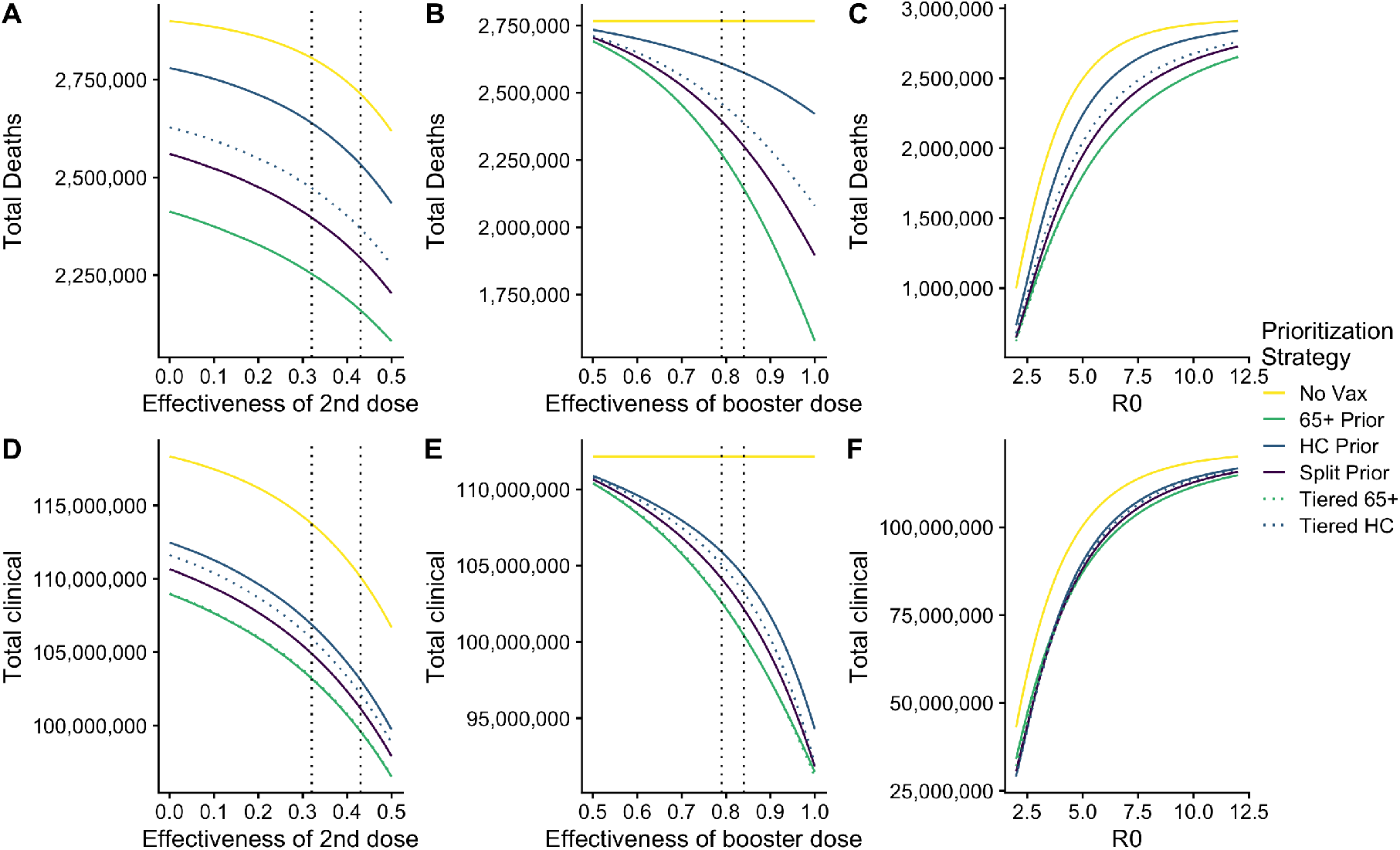
Relationship of stochastically drawn parameters in assessing the effect of booster dose prioritization.

## 4 Discussion

We find that the optimal strategy for reducing deaths due to COVID-19 via vaccination was a tiered roll out that first prioritized adults over age 65 followed by high contact workers, with remaining doses split among low contact adults 18-64 and children (Tiered 65+ strategy). However, the most clinical infections are averted through a tiered roll out that prioritized high contact workers followed by adults over age 65 before general distribution (Tiered HC strategy).

Optimal distribution of booster doses to limit deaths is similar to the primary scenario—it is optimal to prioritize seniors for reducing mortality—and this strategy may also reduce the number of clinical infections the most depending on the transmissibility of the novel variant strain. This difference for booster dose prioritization strategies is driven almost entirely by the lower uptake rate for primary and booster doses by 18-64 year olds compare to adults 65+. However, we note that all three distribution strategies for booster doses result in nearly equal reductions in clinical infections.

These simulations were designed to test the total effect of vaccination—both the direct benefits of vaccination on the prioritized group and the indirect effects of reducing community spread through social contact patterns—using empirical estimates of contact rates across groups. When distributing initial doses, the indirect effects of vaccinating high-risk workers or splitting vaccines between workers and seniors did not confer a greater reduction in deaths than the direct effects of prioritizing all seniors but did reduce the spread of COVID-19 overall.

While limiting deaths is the highest priority, prioritizing high contact workers with in-person employment may alleviate strain on the healthcare system and reduce the number of people living with chronic COVID-19 symptoms. In a real-world setting, there is undoubtedly a benefit to prioritizing some essential workers in healthcare or eldercare settings who have more exposure to high-mortality populations; substantial heterogeneity exists even within our surveyed high risk workers. Results of our ‘split’ analysis show that when some vaccines for adults 65+ are diverted to high risk workers, fewer deaths are averted, but the total number of clinical infections drops sharply. The most lives are saved when adults 65+ are prioritized, which is consistent with early CDC guidance that prioritized the elderly and a limited number of healthcare workers first.

Our results also highlight the significant impact of imperfect vaccines and imperfect vaccine uptake on clinical infections and deaths. We show that the potential disease-averting power of vaccines is lost due to delays in vaccine availability for the general population, as many susceptibles awaiting the vaccine are infected before they are able to be vaccinated. This is especially problematic for strategies prioritizing high-contact individuals as they are more likely to be infected earlier on in the outbreak before vaccination is fully rolled out. Further, low uptake rates, particularly for booster doses can limit the impact of vaccines.

Our study has several limitations. Although our population is faceted by age and employment, the model does not consider other non-pharmaceutical interventions, such as the use of face masks, that can reduce transmission during interpersonal contact. Brief contact between high-risk workers and customers that are masked and distanced may be ultimately insignificant compared to adults lacking such contacts but who are engaging in risky behavior during personal time. Quantifying overall risk along other dimensions of contact, such as whether the contact took place indoors out outdoors, the duration of contact, whether a mask was worn, etc., is an important direction for future research.

This analysis considers prioritization only by age and worker status and does not include heterogeneity within or between groups. Structural barriers in health care, including financial cost, health complications, and discrimination within the healthcare system—if related to essential worker status—may mean that essential workers face increased health risks in addition to increased contact network size.

There are also likely to be benefits to prioritizing people for vaccination along other socio-demographic axes of risk that are not considered here—especially race, ethnicity, and geographic location—by identifying social inequality itself as a driver of outbreaks (Wrigley-Field, Kiang, et al. 2021; Link and Phelan 1995). The BICS survey population reflects the composition of the essential workforce in the United States — Blacks and Hispanics comprise of a larger share of the high-contact population (11.12% and 22.49% respectively) relative to their shares in 65+ population (9.12% and 5.04% respectively). Nonetheless, given the strong age gradient in mortality due to COVID-19, we find that prioritizing the tiered 65+ strategy averts the most deaths among respondents of all ethnicities (see supplement S1.10), assuming that transmission and mortality are equal by ethnicity and gender within our defined demographic groups (children, HC adults, LC adults, adults 65+), we find. Unfortunately, due to limitations in the sample size we are unable to construct a contact matrix that is disaggregated by age, employment, and race or ethnicity, and thus unable to fully account for heterogeneities across these groups. Although the results of our simulations indicate that relying on indirect effects of prioritizing high-contact workers for vaccination does not ultimately save more lives, high-contact workers may be at increased risk of death compared to other adults because of socio-demographic disadvantage within with healthcare system not captured within our model. Our analysis supports prioritization of vaccination for the highest risk members of society—which we identify through advanced age, but may instead be a complex combination of socio-demographic disadvantage. This is an important direction for future research.

Overall our results highlight the impact of two dimensions of risk — contact behavior and age-dependent susceptibility and risk of severe disease— on the effect of vaccination strategies. This work also shows the utility of combining mathematical models with detailed contact data by socio-demographic groups, particularly for understanding sensitivity of the effectiveness of different prioritization strategies to the epidemiology of the circulating virus strain. As novel strains are likely to continue to emerge (Brüssow 2022), these data and models can be especially useful for tailoring prioritization strategies for subsequent outbreaks.

### 4.1 Data availability

We have deposited our data in the Harvard Dataverse, https://doi.org/10.7910/DVN/K8YPVZ (Roubenoff, D. Feehan, and Mahmud 2022a).

### 4.2 Replication Code

All analyses was conducted using R software (version R version 4.0.2). Replication code is publicly available at https://github.com/eroubenoff/BICS_employment_replication_code (Roubenoff, D. Feehan, and Mahmud 2022b).

## Data Availability

4.1 Data availability
We have deposited our data in the Harvard Dataverse, https://doi.org/10.7910/DVN/K8YPVZ (Roubenoff, D. Feehan, and Mahmud 2022a).
4.2 Replication Code
All analyses was conducted using R software (version R version 4.0.2). Replication code is publicly available at https://github.com/eroubenoff/BICS_employment_replication_code (Roubenoff, D. Feehan, and Mahmud 2022b).

## 4.3 Acknowledgements

The authors thank Taylor Chin for her feedback and code sharing. Seed funding was provided by a Berkeley Population Center pilot grant (NICHD P2CHD073964) and further funding was provided by the Hellman Fellows Program.

## Supplementary Material

### S1 Epidemiological Model

#### S1.1 Contact Matrix

**Table S1:** Baseline Contact Matrix ***C*** used in simulations

|  | Child | Adult LC | Adult HC | 65+ |
| --- | --- | --- | --- | --- |
| Child | 1.637 | 0.816 | 1.197 | 0.062 |
| Adult LC | 0.455 | 1.918 | 1.283 | 0.311 |
| Adult HC | 1.285 | 2.472 | 4.550 | 0.495 |
| 65+ | 0.093 | 0.834 | 0.689 | 0.908 |

#### S1.2 Derivation of contacts for the youngest age group

Since the youngest age group (below 18 years of age) were not included in the survey, we use data from the POLYMOD survey (Mossong et al. 2008) in the United Kingdom. First, BICS respondents are re-grouped without employment disaggregation into three age categories—0 − 18,14 − 65, and 65+—and used to derive reciprocal weighted average contact matrix ***B***. Then, data from the POLYMOD survey are grouped into the same three age categories and likewise used to create reciprocal weighted average contact matrix ***P***. As in D. M. Feehan and Mahmud 2021, we calculate the ratio of dominant eigenvalues (*ρ*) of ***B*** and ***P***. The entry for the youngest age group in ***C*** is then taken as the entry for the youngest age group in ***P*** scaled by this ratio:

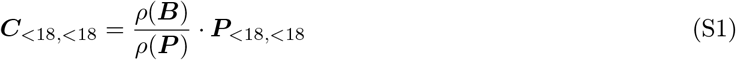

#### S1.3 Alternate Specification of the Contact Matrix

Two alternate specifications of the contact matrix are used for sensitivity tests. First, unlike in the primary simulation where high/low contact status was randomly assigned (in equal proportions to adult respondents surveyed) to alters whose contact status was indeterminable, a matrix is derived without the above described process for unknown alter redistribution. As a number of reported alters are of unknown risk status (no occupation or purpose), in the baseline matrix we assume that the distribution of alters’ contact status at the aggregate level is equal to egos’ contact status. Instead of randomly re-labeling unknown adult alters such that the reported alters have the same survey-weighted proportion of high/low contact status as egos, in this alternate specification we label all unknown alters as having Low Contact status. This represents a conservative estimate for alter contact patterns. Second, we derive the contact matrix otherwise as normal, but with an alternate specification for the POLYMOD matrix ***P*** used to derive child-child contacts. This departs from the baseline simulation, where we use POLYMOD contacts with school contacts removed to represent school closures at the time of writing. For this alternate specification, we derive the matrix that includes these POLYMOD contacts.

The primary simulation is run with these two alternate contact matrix specifications contact matrix (shown in figure S1). All other parameters are as baseline. Both total deaths and clinical infections are consistently higher with the two alternate specifications compared to baseline. In neither set of simulations did the the optimal strategies change.

**Table S2:** Contact Matrix without random unknown alter reallocation

|  | Child | Adult LC | Adult HC | 65+ |
| --- | --- | --- | --- | --- |
| Child | 1.637 | 0.858 | 1.207 | 0.062 |
| Adult LC | 0.478 | 2.009 | 1.260 | 0.319 |
| Adult HC | 1.296 | 2.427 | 4.332 | 0.486 |
| 65+ | 0.093 | 0.855 | 0.677 | 0.922 |

**Table S3:** Contact Matrix derived without POLYMOD school contacts removed

|  | Child | Adult LC | Adult HC | 65+ |
| --- | --- | --- | --- | --- |
| Child | 3.271 | 0.847 | 1.193 | 0.060 |
| Adult LC | 0.472 | 1.930 | 1.277 | 0.304 |
| Adult HC | 1.281 | 2.459 | 4.491 | 0.524 |
| 65+ | 0.090 | 0.814 | 0.730 | 0.874 |

**Figure S1:**
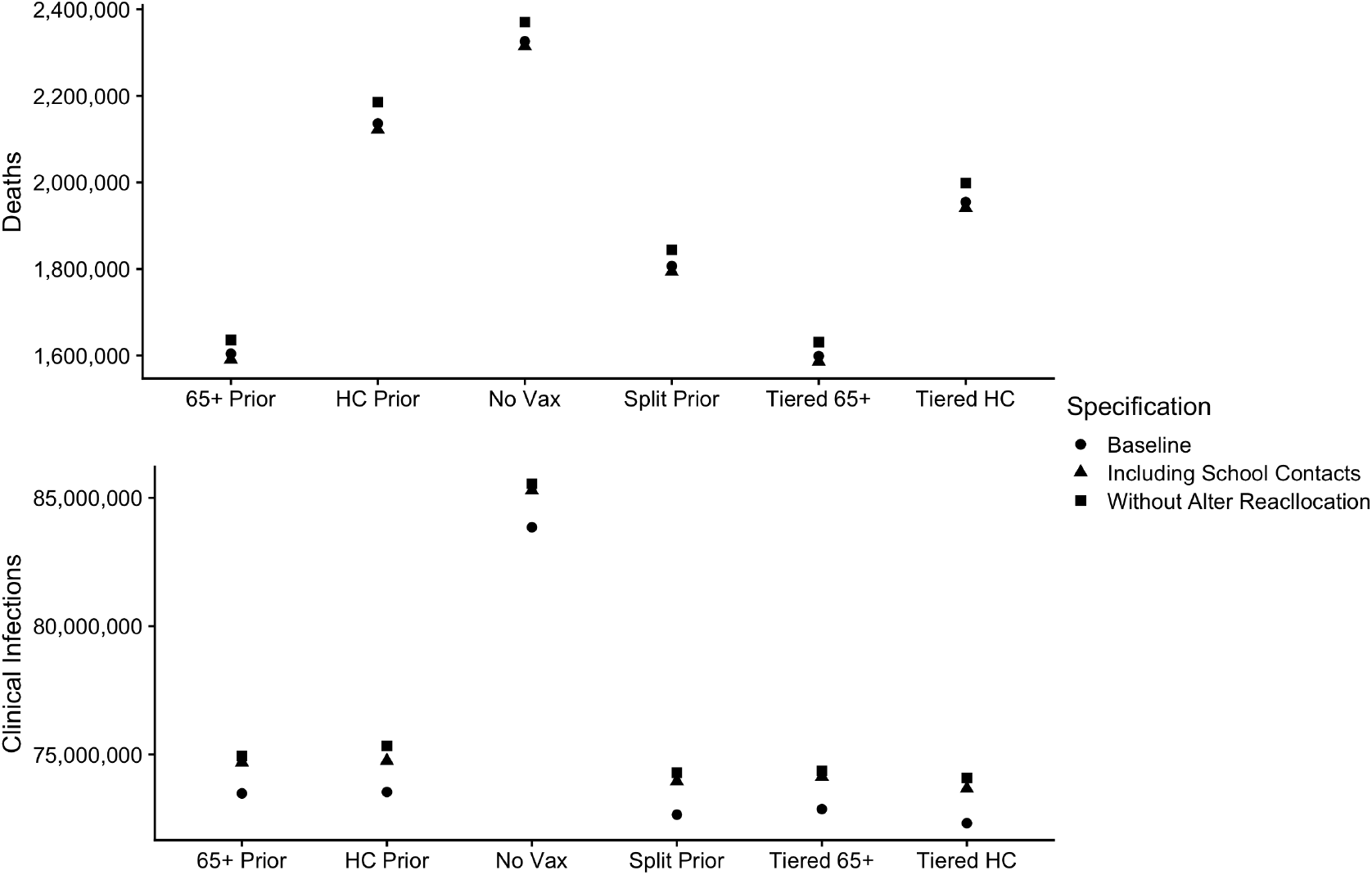
Deaths and infections for the 5 intervention strategies compared to null for alternate contact matrix specifications, without random unknown alter reallocation and without POLYMOD school contacts removed.

#### S1.4 Derivation of the Starting Population

The starting population is derived from the Johns Hopkins COVID-19 dashboard data repository (JHU; Dong, Du, and Gardner 2020) and American Community Survey 2019 estimates (ACS; US Census Bureau 2019) using parameters from the baseline simulation listed in table S8. The JHU data contain the state level counts of confirmed COVID-19 cases and deaths; in addition, 45 of 51 states (including DC) list counts of active cases. To obtain the initial size of the clinically infectious compartment, we scaled the national count of total confirmed cases by the proportion of cases currently active for states with active cases listed. For these 45 states with listed active cases, on 1 January 2021 there were 14, 405, 003 total confirmed cases, of which 40.5% (5, 829, 986) were currently active. This proportion is used to scale the total confirmed cases (20, 278, 813) to yield the total initial size of the clinically infectious compartment: 8, 207, 366.

The BICS wave 4 survey indicated that, adjusted for survey weights, 34% of adults were working in person (HR) and 66% were not (LR; including unemployed). These survey data were used to scale the ACS age-disaggregated data to yield the total group sizes N:

**Table S4:** Group sizes N derived from the ACS

| Child | LC Adult | HC Adult | 65+ |
| --- | --- | --- | --- |
| 73,429,392 | 132,319,841 | 68,164,766 | 50,783,796 |

We assume that the total count of Active cases is distributed proportionally to group size (N) and the age-dependent probability of infection (u, table S8) to produce infectious and symptomatic compartment *Ic*(0):

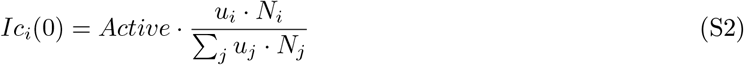

Individuals in the exposed (pre-infectious) compartment are taken to be proportional *I*_*c*_. The constant of proportionality is taken as the ratio of average time spent in the *E* compartment to average time spent in the *I*_*c*_ compartment. This assumes that the size of the outbreak is stationary at the start of the simulation.

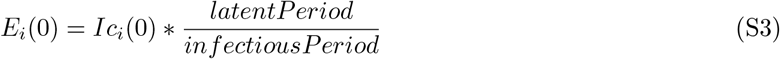

Remaining members of the population are all assumed to be susceptible. Susceptibles are then divided into *S* and *S*_*x*_, indicating if they are awaiting or declining the vaccine. We assume that all groups will have 70% uptake. For baseline parameters of *μ* and *ρ*, the initial population state is:

**Table S5:** Primary Simulation Starting Population

|  | Child | Adult LC | Adult HC | 65+ |
| --- | --- | --- | --- | --- |
| $N$ | 73,553,240 | 132,073,398 | 68,037,811 | 49,238,581 |
| $S$ | 50,346,846 | 88,093,322 | 45,381,408 | 33,018,446 |
| $S_x$ | 21,577,220 | 37,754,281 | 19,449,175 | 14,150,763 |
| $E$ | 610,941 | 2,334,673 | 1,202,710 | 776,015 |
| $I_c$ | 1,018,235 | 3,891,122 | 2,004,517 | 1,293,358 |
| $I_{sc}$ | 0 | 0 | 0 | 0 |
| $R$ | 0 | 0 | 0 | 0 |
| $D$ | 0 | 0 | 0 | 0 |
| $V_a$ | 0 | 0 | 0 | 0 |
| $V_b$ | 0 | 0 | 0 | 0 |
| $V_{bx}$ | 0 | 0 | 0 | 0 |
| $V_{boost}$ | 0 | 0 | 0 | 0 |

In the booster simulation, we utilize uptake data derived from the CDC to initialize a proportion of the susceptible population in compartment *V*_*b*_, indicating that they have received a primary course of vaccination. Individuals who decline the primary vaccine remain in compartment *S*_*x*_ until infected. We assume that 70% of individuals who have received a primary course of vaccination will receive boosters; the remaining 30% are placed in compartment *V*_*bx*_, indicating that they decline the vaccine.

Finally, we conducted a series of simulations assuming high uptake of the primary vaccine among both low and high contact adults to match older adult uptake (95%). These simulations closely mirrored those for the main booster simulations—prioritizing older adults, either solely or in a tiered roll-out limited the most deaths, and the number of infections limited was nearly equal across all strategies.

**Table S6:** Booster Simulation Starting Population

|  | Child | Adult LC | Adult HC | 65+ |
| --- | --- | --- | --- | --- |
| $N$ | 73,553,240 | 132,073,398 | 68,037,811 | 49,238,581 |
| $S$ | 0 | 0 | 0 | 0 |
| $S_x$ | 47,462,186 | 27,092,608 | 13,956,798 | 2,394,698 |
| $E$ | 615,314 | 2,351,386 | 1,211,320 | 781,570 |
| $I_c$ | 1,025,523 | 3,918,976 | 2,018,866 | 1,302,616 |
| $I_{sc}$ | 0 | 0 | 0 | 0 |
| $R$ | 0 | 0 | 0 | 0 |
| $D$ | 0 | 0 | 0 | 0 |
| $V_a$ | 0 | 0 | 0 | 0 |
| $V_b$ | 17,115,152 | 69,097,300 | 35,595,579 | 31,331,788 |
| $V_{bx}$ | 7,335,065 | 29,613,129 | 15,255,248 | 13,427,909 |
| $V_{boost}$ | 0 | 0 | 0 | 0 |

**Table S7:**
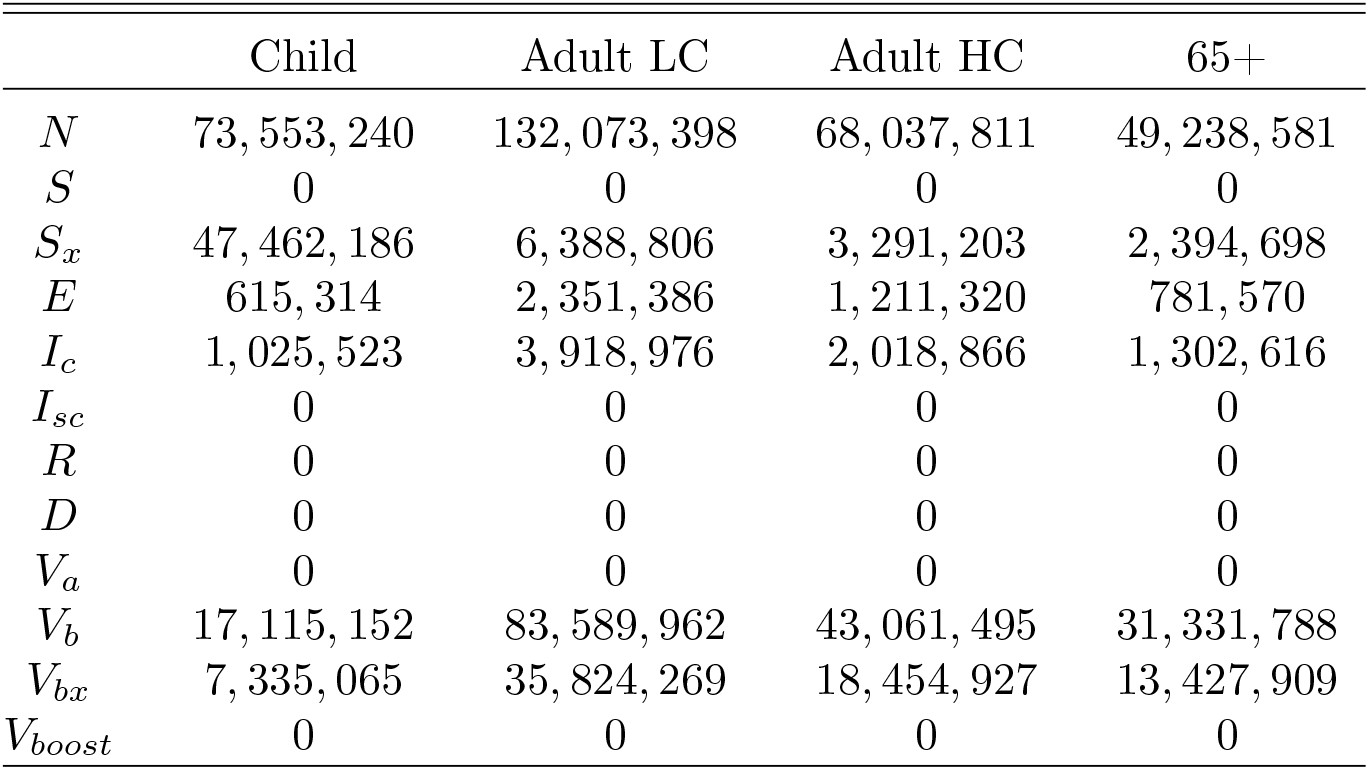
Booster Simulation Starting Population with High Primary Uptake

#### S1.5 Primary (2-dose) vaccination model

**Figure S2:**
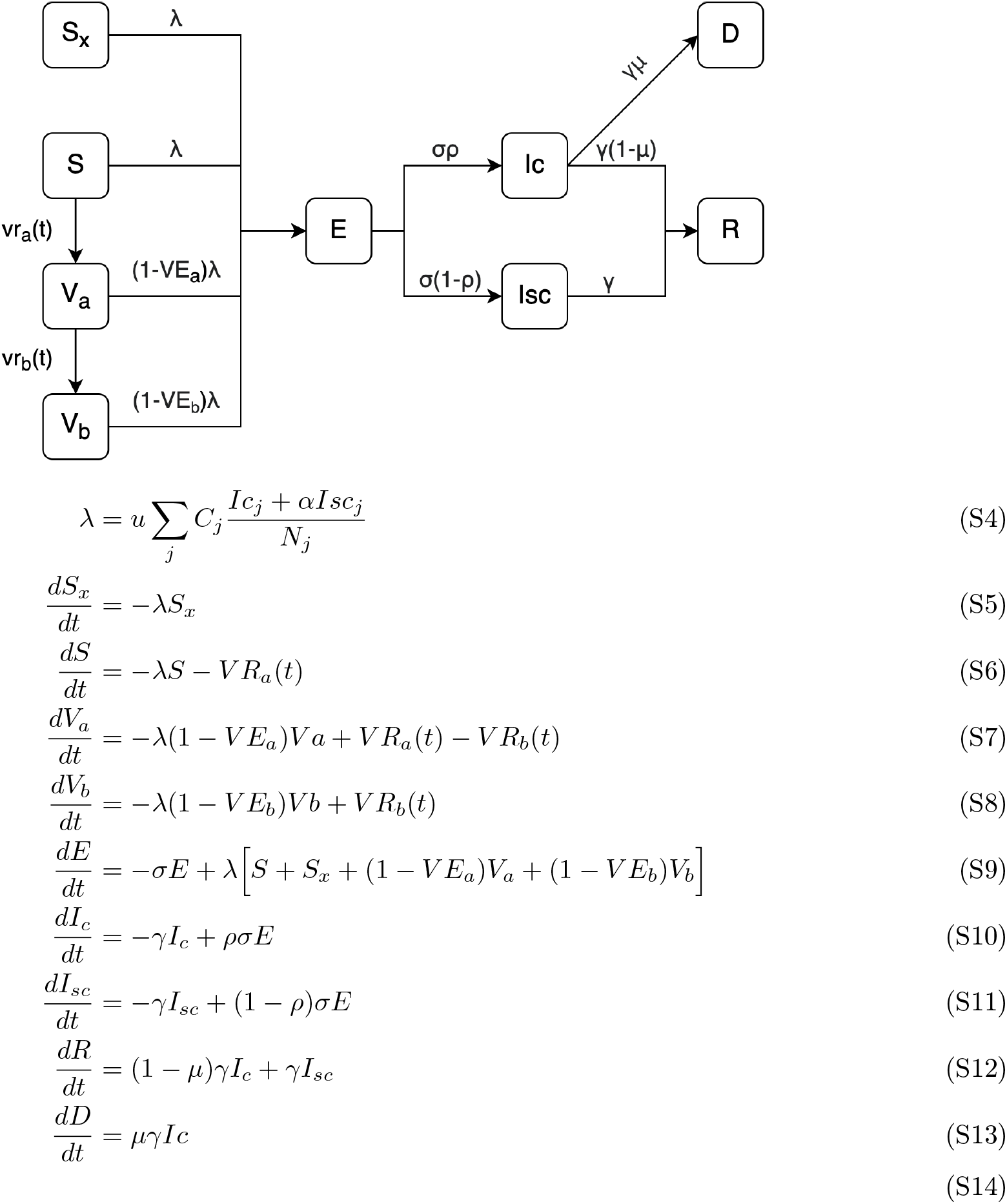
Primary vaccination model. Compartments *S, S*_*x*_, *E, I*_*c*_, *I*_*sc*_, *R, D, V*_*a*_, *V*_*b*_ correspond to susceptible and waiting to be vaccinated, susceptible but not waiting to be vaccinated, exposed, clinically infectious, subclinically infectious, recovered, deceased, vaccinated 1st dose, and vaccinated 2nd dose, respectfully. Parameter *u* is the group-specific probability of infection upon contact with an infected person; *C*_*ij*_ is the number of contacts a person in group *i* has with a person in group *j*; *α* is the relative infectiousness of subclinical cases; *V E*_*a,b*_ are reduction in infection after 1 and 2 doses of the vaccine; *ρ*_*i*_ is probability of symptomatic illness after exposure and first and second shots; *μ, σ, γ* are the mortality, rate of progressing from *E* to I, and recovery rate. *V R*_*ai,bi*_(*t*) are the time-specific vaccination rates for 1st and 2nd vaccines. *NB: Group index i is dropped to reduce visual clutter*.

#### S1.6 Booster (3rd dose) vaccination Model

**Figure S3:**
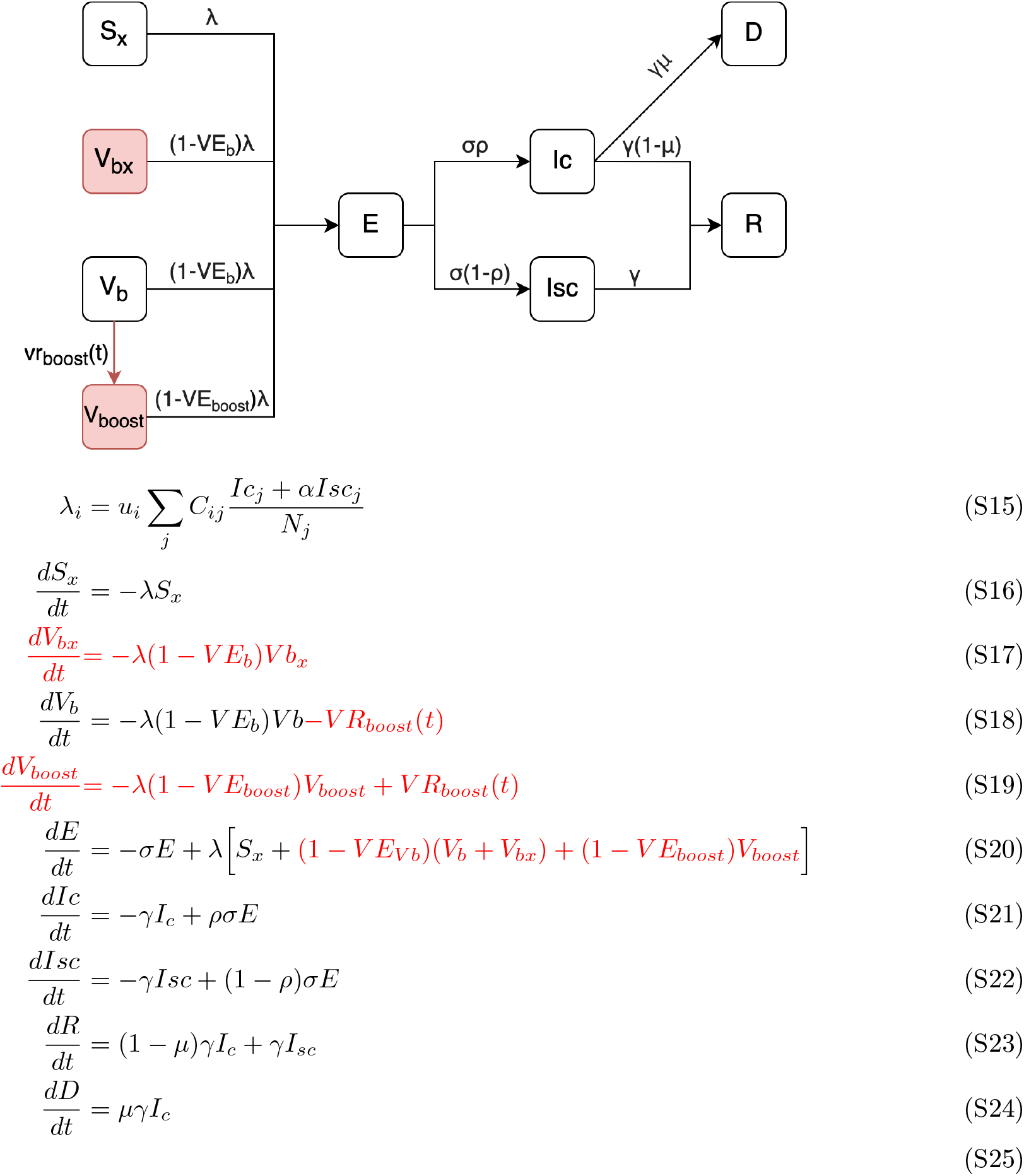
The model is extended to accommodate the prioritization of additional booster doses for people who have already received two doses of the vaccine (compartment *V*_*b*_). Additions to the model are indicated in red; compartments *S* and *V*_*a*_ are removed as no shots are allocated to unvaccinated individuals. Susceptible compartments *S*_*x*_, *V*_*bx*_, *V*_*b*_, and Vboost indicate resepectfully to individuals who have received no doses of the vaccine but will not receive boosters, those who have received two doses but will not get booster doses, those waiting for booster doses, and those who have received booster doses. *V Rboost*_*i*_ is the rate at which booster doses are given to people in *V b*_*i*_. *V E*_*b,boost*_ indicate the efficacy of the vaccine at reducing transmission. *NB: Group index i is dropped to reduce visual clutter*.

#### S1.7 Vaccine distribution

We took a general value of 2 million vaccine doses distributed daily in all primary simulations. There are six strategies tested for a set of parameters:

- No Vax: No vaccination (no priority or general doses distributed)
- 65+ Prior: Prioritizing only seniors before general distribution
- HC Prior: Prioritizing only adults 18-64 with in-person contacts before general distribution
- Split Prior: Splitting priority vaccines evenly (i.e., 500,000 each) between seniors and working adults before general distribution
- Tiered 65+: ‘Tiered’ rollout that prioritizes, in order, seniors before working adults before general distribution
- Tiered HC: ‘Tiered’ rollout that prioritizes, in order, working adults before seniors before general distribution

Here, ‘general distribution’ refers to distributing vaccine doses proportionally to remaining group size. For all strategies besides the no vaccination baseline, general distribution occurs after all priority doses have been distributed. The tiered strategies are intended to replicate the CDC’s guidance, which opened up priority eligibility to the elderly, high risk people, and essential workers in progressive tiers before opening up eligibility to all. In the primary 2-dose vaccination model, the 2 million eligible doses are split evenly between first and second doses, meaning that 1 million susceptible can receive the first dose daily and 1 million people in *V*_*a*_ can receive the second dose daily.

#### S1.8 Model Parameters

**Table S8:** Description of parameters for baseline model. Groups specified are for children, all adults, and seniors, respectively.

| Parameter | Description | Value | Source |
| --- | --- | --- | --- |
| $C_{ij}$ | Contact rate between individuals in group $i$ and $j$ | Estimated | Berkeley Interpersonal Contact Survey (BICS) |
| $\alpha$ | Relative Infectiousness of subclinical and clinical cases | 0.5 | Assumption, in line with the range presented in McEvoy et al. 2020 |
| $1/\sigma$ | Mean latent period | 3 days | Davies et al. 2020; Bubar et al. 2021 |
| $1/\gamma$ | Mean infectiousness period | 6 days | Davies et al. 2020; Bubar et al. 2021 |
| $\rho$ | Probability of having clinical symptoms per age group | [0.26, 0.36, 0.69] | Davies et al. 2020 |
| $u$ | Relative susceptibility to infection per age group | [0.39, 0.83, 0.74] | Davies et al. 2020; Bubar et al. 2021 |
| $\mu$ | Mortality per group | [0.004%, 0.023%, 8%] | Levin et al. 2020 |
| $R_0$ | Basic Reproduction Ratio | 2.5 | D. M. Feehan and Mahmud 2021 |
| $VE_a,$<br>$VE_b$ | Primary model: Vaccine effectiveness after first dose and second dose | 0.80, 0.90 | Tenforde 2021; Thompson 2021 |
| $VE_b,$<br>$VE_{boost}$ | Booster model: Vaccine effectiveness after second dose and booster dose | 37%, 81% | Thompson 2022 |
| $v_t$ | Time between first and second primary doses | 25 days | Assumed |

The force of infection *λ*_*i*_ for a susceptible individual in group *i* is:

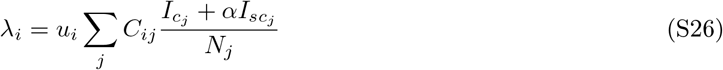

where *u*_*i*_ is the age-dependent susceptibility to infection after contact with an infectious individual, *C*_*ij*_ is the entry in contact matrix ***C*** corresponding to the average number of daily contacts a respondent in group *i* has with an individual in group *j*; *I*_*c*_*j* and and *I*_*sc*_*j* are the number of clinically infectious and subclinically infectious individuals in group *j, α* is the relative infectiousness of clinical vs. subclinical infections, and *N*_*j*_ is the total population size (Davies et al. 2020; Bubar et al. 2021).

**Table S9:** Summary of the 5 vaccination prioritization scenarios for analysis

| Strategy | Description |
| --- | --- |
| No Vax | Null scenario; no vaccine distribution |
| 65+ Prior | Initial prioritization for seniors over age 65; remaining vaccines distributed proportionally to eligible group size |
| HC Prior | Initial prioritization for high contact workers; remaining vaccines distributed proportionally to eligible group size |
| Split Prior | Priority vaccine dose are split evenly between seniors over age 65 and high contact workers; remaining vaccines distributed proportionally to eligible group size |
| Tiered 65+ | First priority vaccines are given to seniors over age 65 and second to high contact workers; remaining doses are distributed to children and low contact adults proportional to eligible group size |
| Tiered HC | First priority vaccines are given to to high contact workers and second to seniors over age 65; remaining doses are distributed to children and low contact adults proportional to eligible group size |

#### S1.9 Booster Model Sensitivity Analysis

We find that the optimal prioritization strategy is highly sensitive to model parameters (figure 6), suggesting that the epidemiology of any subsequent variants will determine the optimal vaccination strategy for booster doses. Holding all other parameters at the baseline median value, we varied the efficacy of the primary course of vaccination from 0 to 50%, efficacy of the booster doses between 50% and 100%, and *R*_0_ between 2 and 12. All other parameters are held at baseline values. We find that the reduction in deaths is nearly always higher under senior prioritization than under high risk adult prioritization. Efficacy of the primary and booster doses plays a substantial role in shaping the outbreak; 100% effective booster doses reduce deaths by nearly 50% under 65+ prioritization, whereas a 50% effective booster only reduces deaths by 10%. As *R*_0_ increases from 2 to 12, we observe a similar cross-over in clinical infections as the primary scenario: at values of *R*_0_ below 3.6, Tiered HC and HC Prioritization limits the most clinical infections; above this value, 65+ and Tiered 65+ Prioritization limit the most clinical infections. Additionally, higher values of *R*_0_ generally correspond to less effective interventions, since the outbreak peak is higher by the time of widespread booster uptake.

Investigation of clinical infections averted during High Contact adult booster prioritization (figure 5B) reveals an interesting dynamic where counts of averted infections relative to the no booster scenario appear to stall after mid-January. Further investigation of group-level incidence rates (supplemental figure S9) reveals that when this group is prioritized in part or in full, after mid January counts of infections among HC adults are actually lower in the null scenario—where no booster doses are distributed—than simulations that included booster distribution. We observe that the HC adult curve is ‘flattened,’ where the peak of the curve is lowered but infections continue to occur for longer. Ultimately only about 500,000 fewer clinical infections occur during HC Prioritization (of about 23 million in the null scenario), indicating that this strategy is much less effective at high values of *R*_0_.

Uptake of the primary vaccine is considerably lower for adults between ages 18 and 65 and than for seniors; as a result, proportionally fewer working-age adults are eligible for a booster dose than seniors. As a sensitivity test, we ran an additional 1000 simulations assuming that HC and LC adult primary vaccine uptake rate matches the 65+ adult uptake rate of 95%; booster uptake remains at 70% of the eligible primary-vaccinated population. 1000 simulations are conducted with randomly drawn *R*_0_ and vaccine efficacy parameters identical to the above simulation. We find a similar overall story: prioritizing seniors first, either solely or in a tiered roll-out, limits the most deaths, and the reductions in clinical infections is nearly equal between all five strategies. With baseline parameters, tiered 65+ roll-out reduces deaths by 26% compared to no booster doses and tiered HC roll-out reducing clinical infections by 12%. Results are further elaborated in the model supplement.

#### S1.10 Gender and Ethnicity

Table S10 shows the gender and ethnicity of respondents in each category (Adult LC, Adult HC, 65+). High Contact adults are almost 20 percentage points less likely to be female than Low Contact adults. The ethnic breakdown of High Contact adults is similar to that of Low Contact adults. Adults aged 65+ are more about 10 percentage points more likely to be white and 15 percentage points less likely to be Hispanic.

Assuming that contact rates, probability of transmission, and mortality are equal by gender and ethnicity, prioritizing the oldest age group for vaccination averts the most deaths in all demographic categories. Even though Hispanics make up 20% of the adult population but only 5% of the 65+ population, prioritizing the oldest age group still averts nearly 20,000 more Hispanic deaths (about 20%) compared to HC prioritization. Since mortality among the oldest age group is about 350 times higher than for adults ages 18-64, the direct benefit of preventing deaths among this age group through vaccination remains the optimal strategy for vaccine distribution.

In aggregate, Tiered HC vaccine rollout limits the most clinical infections, and this is remains true for Blacks and Asians. HC prioritization (without tiered roll-out) averts the most deaths among Hispanics. However, Tiered 65+ roll-out averts the most infections among Women and Whites, whose age distributions skew slightly older than the other groups. Like in the primary analysis, we note that differences in clinical infections between strategies is small.

**Table S10:** Survey-weighted estimates of Gender and Ethnicity of BICS respondents, by category.

| Category | Female | White | Black | Asian | Hispanic |
| --- | --- | --- | --- | --- | --- |
| Adult LC | 57.68% | 73.75% | 13.56% | 6.04% | 20.56% |
| Adult HC | 38.45% | 76.38% | 11.12% | 4.91% | 22.49% |
| 65+ | 52.49% | 85.27% | 9.12% | 3.29% | 5.04% |

**Figure S4:**
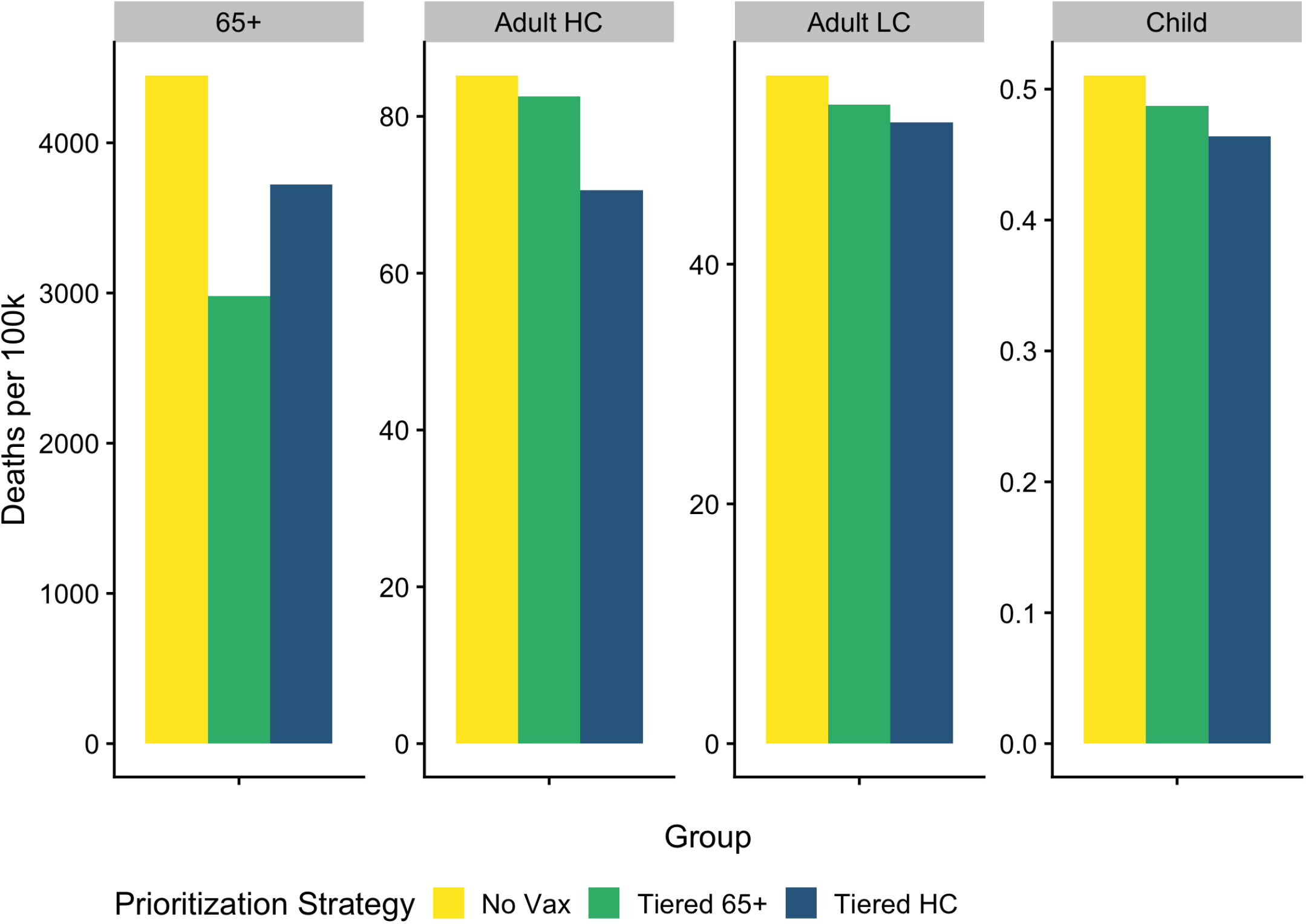
Deaths by category used in transmission model for selected prioritization strategies (chosen to reduce visual clutter), per 100k in each category.

**Figure S5:**
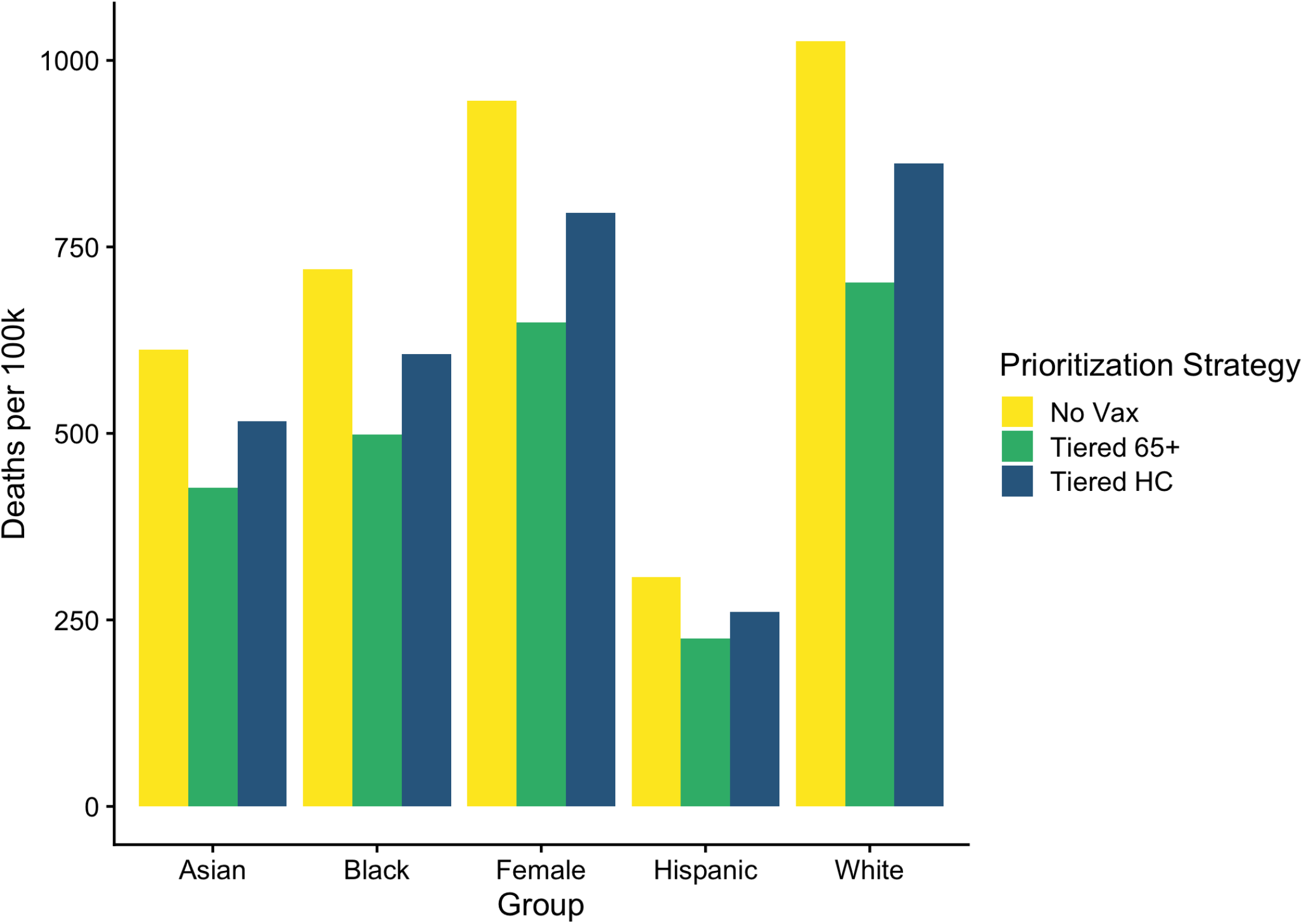
Deaths per 100k by gender and ethnicity, assuming equal transmission and mortality parameters between groups, for selected prioritization strategies (chosen to reduce visual clutter).

### S2 Additional Figures

**Figure S6:**
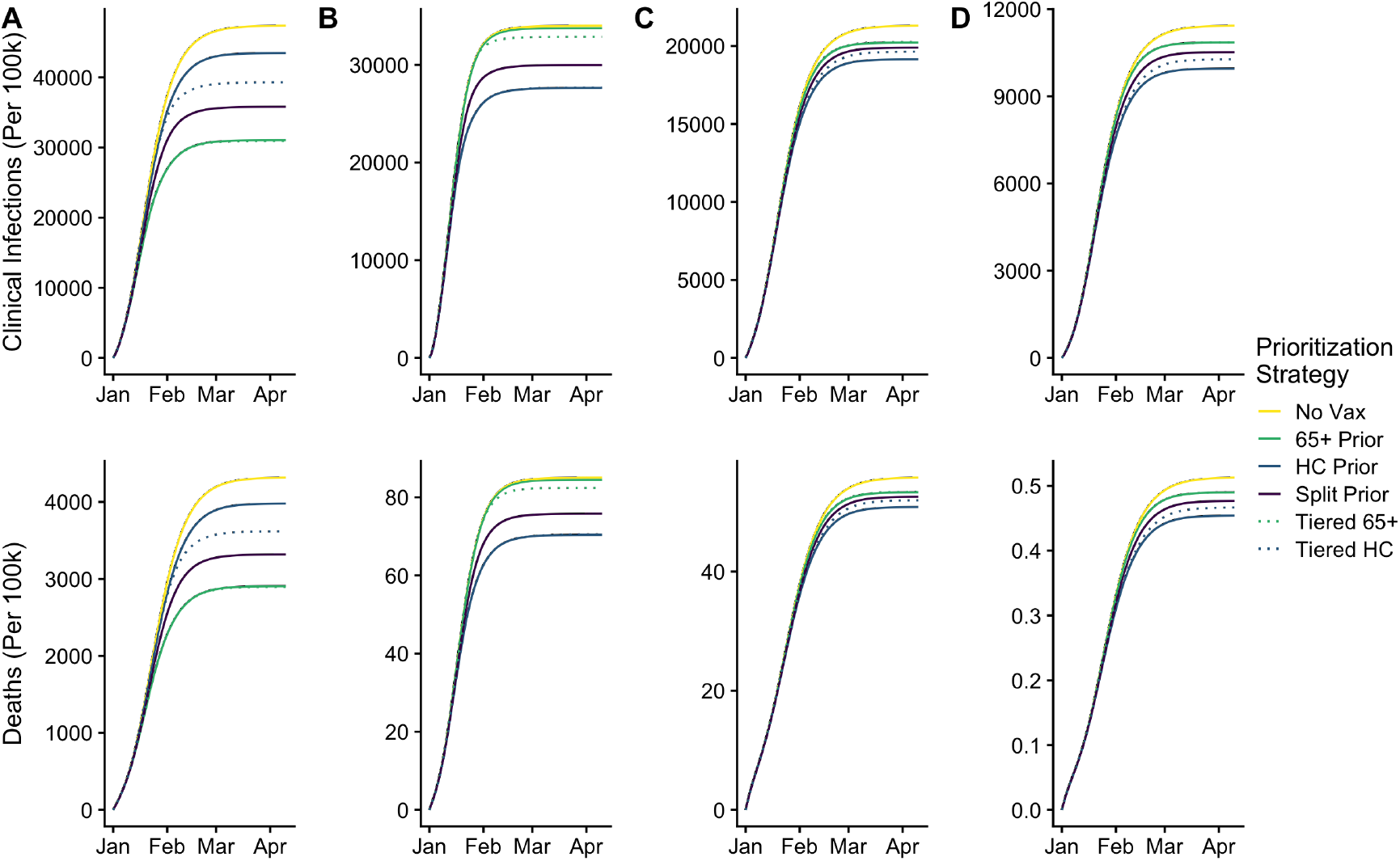
Cumulative clinical infections and deaths during the baseline simulation for all demographic groups (A-D: Seniors, HC Workers, LC Workers, Children) and prioritization strategies per 100,000 individuals in each group. The largest reduction in infections occur from the direct effects of vaccinating HR workers and seniors, but indirect benefits are observed when seniors or workers receive non-priority access. Mortality reductions are greatest when a seniors or HC workers are directly given priority access.

**Figure S7:**
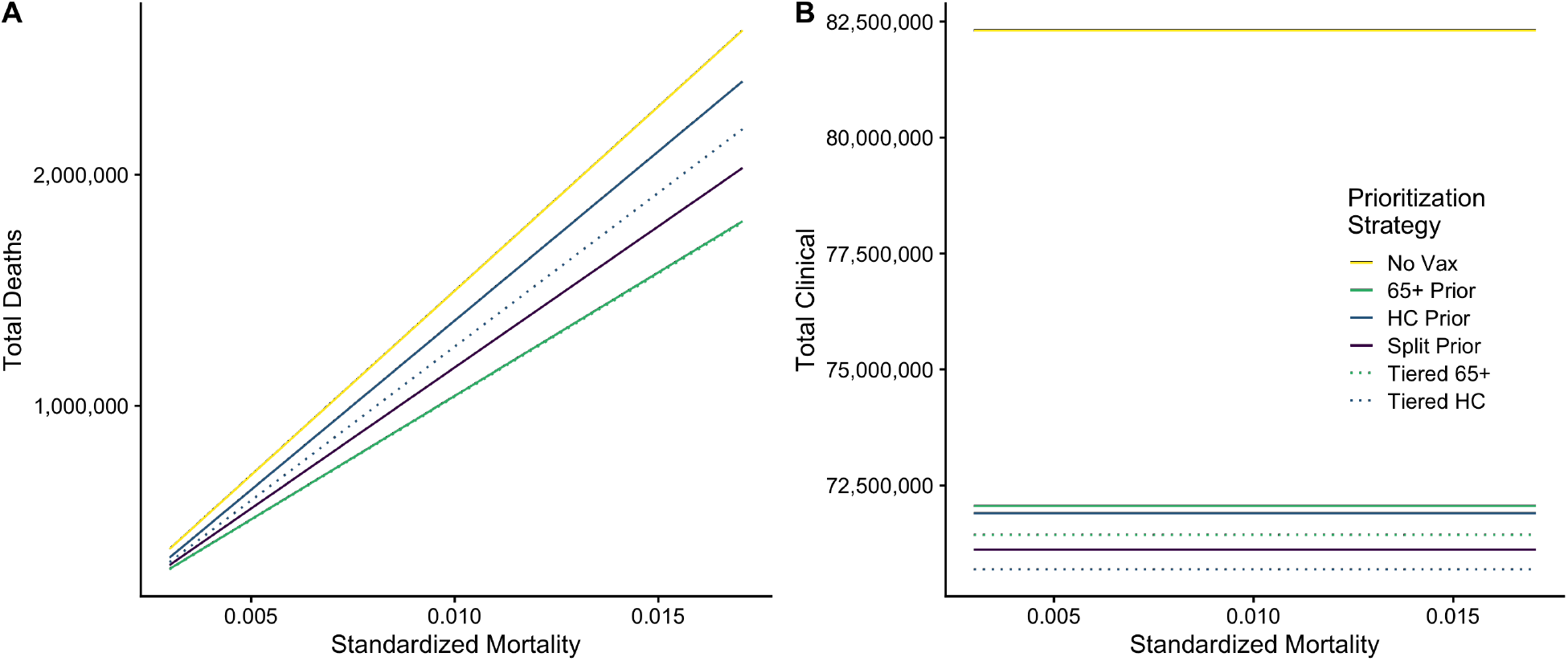
Results of the sensitivity analysis of 65+ mortality *μ*_65+_ (varied between 0.01 and 0.1) and the effect on total deaths (A) and clinical infections (B). X-axis indicates total mortality standardized for group size.

**Figure S8:**
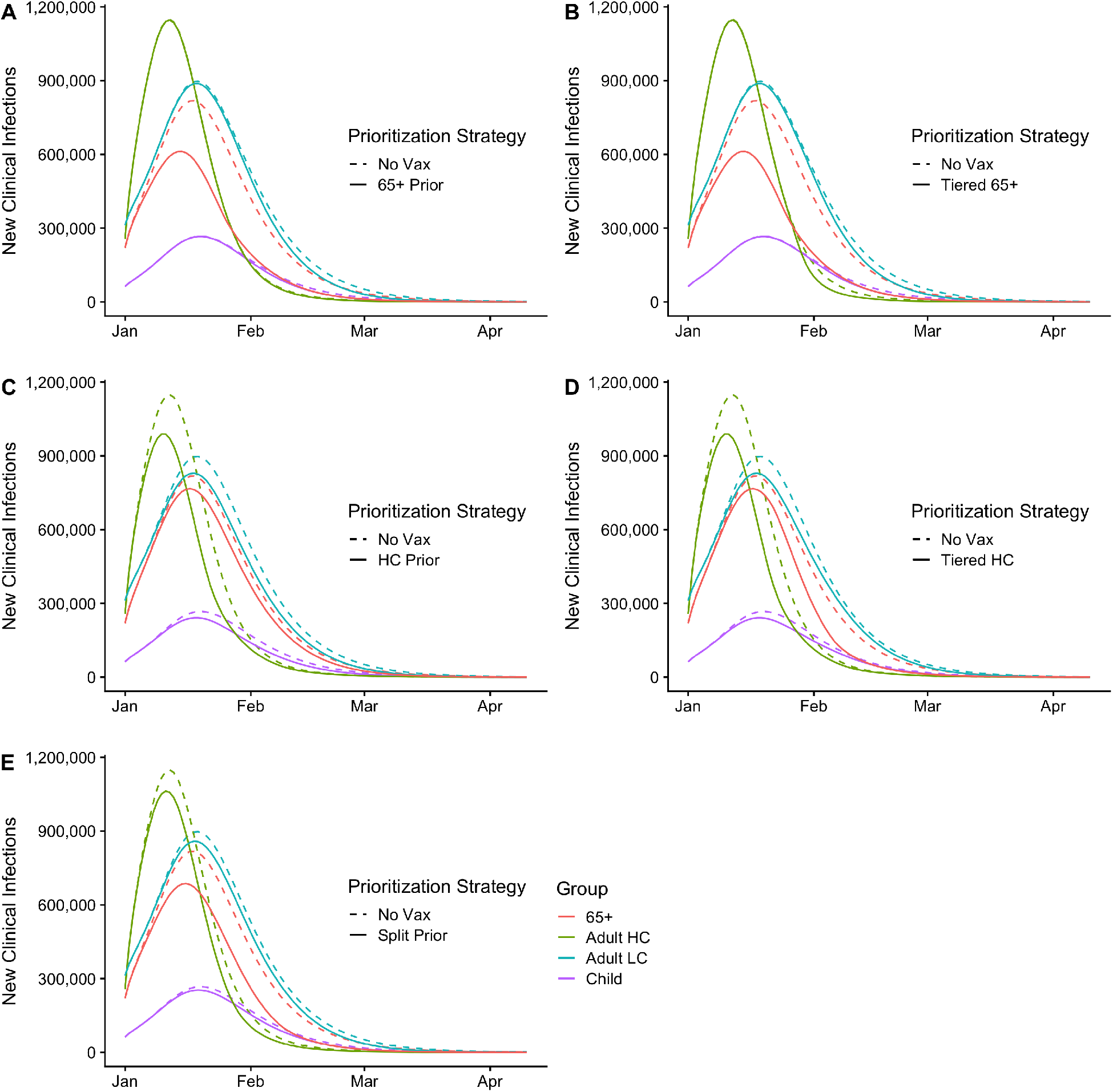
New clinical infections daily by group and prioritization strategy for the primary (2-dose) vaccine distribution model with baseline parameters, with No Vaccine scenario shown as comparison. (A) 65+ Prioritization; (B) Tiered 65+ Prioritization; (C) High Contact Prioritization; (D) Tiered High Contact Prioritization; (E) Split prioritization.

**Figure S9:**
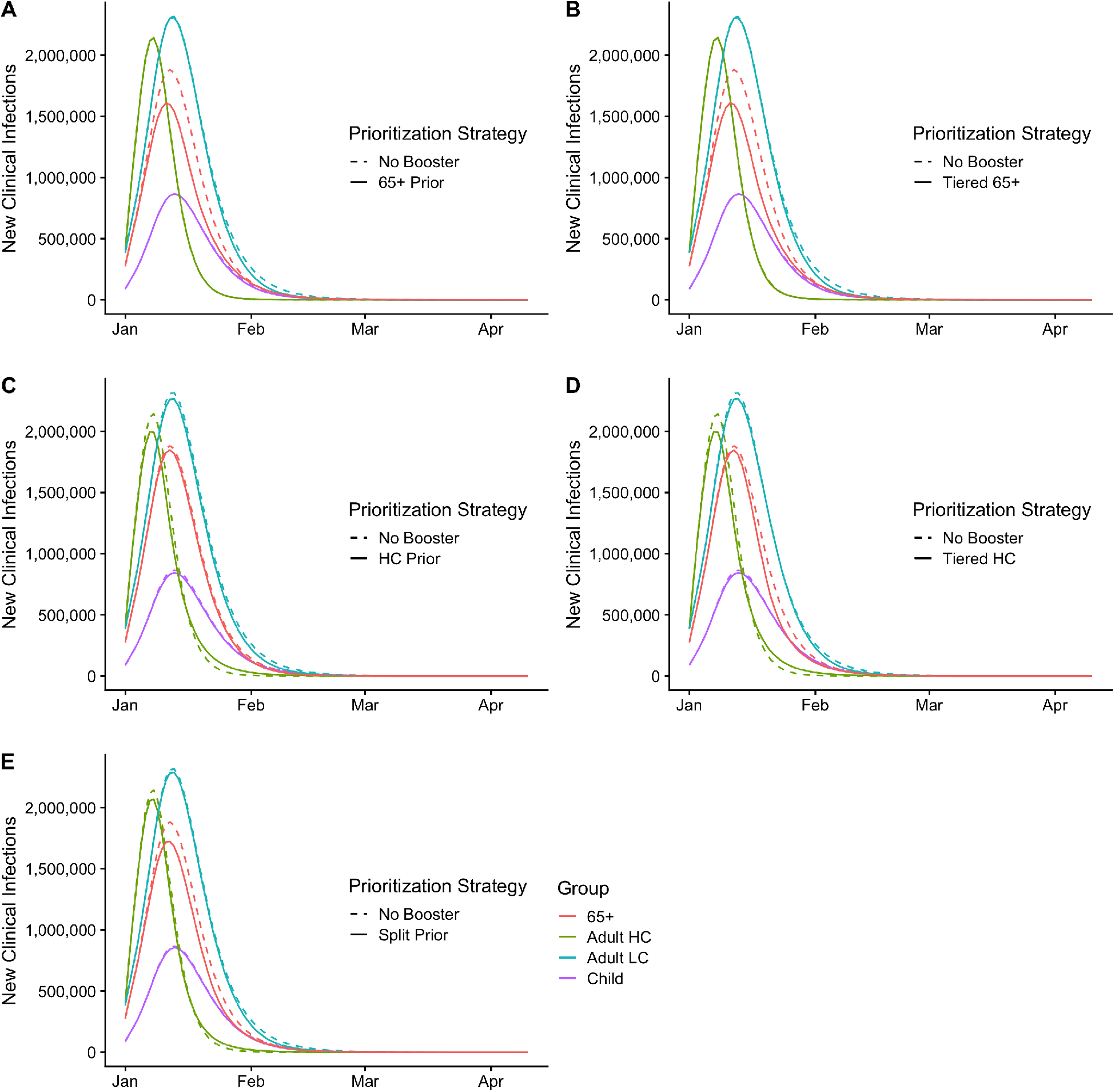
New clinical infections daily by group and prioritization strategy for the booster vaccine distribution model with baseline parameters, with No Vaccine scenario as comparison. (A) 65+ Prioritization; (B) Tiered 65+ Prioritization; (C) High Contact Prioritization; (D) Tiered High Contact Prioritization; (E) Split prioritization. In scenarios where high contact workers receive priority vaccines, either directly, tiered, or split (panels C-D), incidence among that group is lower after the peak of the outbreak during the No Booster scenario than the booster scenario.

**Figure S10:**
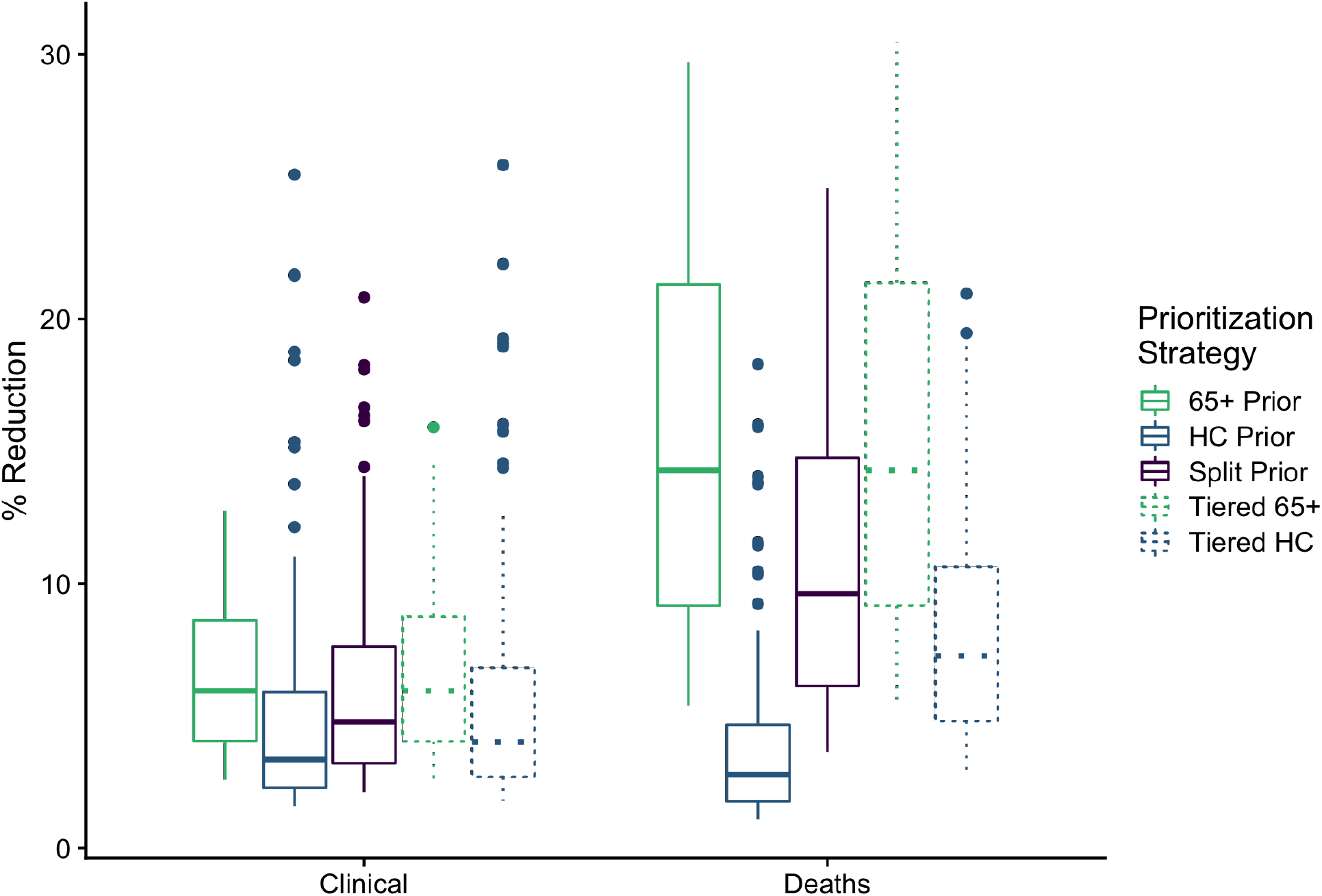
Results from stochastically drawn parameter for high-uptake booster scenario

